# An Integrative Polygenic and Epigenetic Risk Score for Overweight-Related Hypertension in Chinese Population

**DOI:** 10.1101/2025.01.13.25320495

**Authors:** Yaning Zhang, Qiwen Zheng, Qili Qian, Na Yuan, Tianzi Liu, Xingjian Gao, Xiu Fan, Youkun Bi, Guangju Ji, Peilin Jia, Sijia Wang, Fan Liu, Changqing Zeng

## Abstract

Overweight-related hypertension (OrH), defined by the coexistence of excess body weight and hypertension (HTN), is an increasing health concern elevating cardiovascular disease risks. This study evaluates the prediction performance of polygenic risk scores (PRS) and methylation risk scores (MRS) for OrH in 7,605 Chinese participants from two cohorts: the Chinese Academy of Sciences (CAS) and the National Survey of Physical Traits (NSPT). In CAS cohort, which predominantly consists of academics, males showed significantly higher prevalence of obesity, hypertension (HTN), and OrH, along with worse metabolic syndrome indicators, compared to females. This disparity was less pronounced in NSPT cohort and in broader Chinese studies. Among ten PRS methods, PRScsx was the most effective, enhancing prediction accuracy for obesity (AUC = 0.75), HTN (AUC = 0.74), and OrH (AUC = 0.75), compared to baseline models using only age and sex (AUC = 0.55–0.71). Similarly, Lasso-based MRS models improved prediction accuracies for obesity (AUC = 0.70), HTN (AUC = 0.73), and OrH (AUC = 0.78). Combining PRS and MRS further boosted prediction accuracy to the AUC of 0.77, 0.76, and 0.80, respectively. These models stratified individuals into high (> 0.6) or low (< 0.1) risk categories, covering 59.95% for obesity, 31.75% for HTN, and 43.89% for OrH, respectively. Our findings highlight a higher OrH risk among male academics, emphasize the influence of metabolic and lifestyle factors on MRS predictions, and highlight the value of multi-omics approaches in enhancing risk stratification.

**Highlights:** Polygenic risk scores and methylation risk scores were systematically evaluated in predicting the risk of obesity, hypertension, and overweight related hypertension in Chinese participants. PRScsx demonstrated robust accuracy in PRS profiling, while Lasso-based MRS showed superior performance in MRS profiling. Moreover, integrating multi-omics analyses further improved disease risk profiling for these conditions, highlighting their potential for personalized care and prevention strategies.

Gender disparity in the prevalence of metabolism-related disorders largely changed in recent three decades in China. Male to female prevalence ratio for obesity, hypertension, and overweight related hypertension reached striking high as 3.8, 2.9 and 4.7 among academics. These differences are likely influenced by the complex interplay among epigenetic factors, lifestyle and metabolic health.

## Introduction

Overweight-related hypertension (OrH) is a distinct clinical condition characterized by the concurrent disorders of both body weight and blood pressure [1–6]. Its global prevalence has largely increased over the past two decades, linking to a rise in the risks of cardiovascular and cerebrovascular diseases [1, 2, 7, 8]. Recent Genome-Wide Association Studies (GWAS) [9] and Epigenome-Wide Association Studies (EWAS) [10] have uncovered numerous genetic and epigenetic factors associated with body weight and blood pressure. To date, the NHGRI-EBI GWAS catalog [11, 12] has cataloged 4,263 SNPs from 54 studies that are significantly associated with body mass index (BMI), spanning 1,252 genes. Meanwhile, 36 studies have identified 2,853 SNPs across 862 genes significantly associated with diastolic blood pressure (DBP) and systolic blood pressure (SBP) (**Table S1**). Furthermore, EWAS have identified 1,581 CpG sites across 855 genes that are associated with BMI [13–18] along with 150 CpG sites from 85 genes associated with blood pressure (**Table S2**) [19–25].

With the continuous discovery of a large number of genetic and epigenetic risk factors, polygenic risk scores (PRS) [26, 27] and methylation risk scores (MRS) [21, 28, 29] have emerged as pivotal tools for profiling the risk landscape of OrH. For instance, a meta analysis involving 700,000 European individuals constructed a PRS using 941 SNPs, which explained approximately 6% of the variance in BMI [30]. A stratification analysis from the Korean Genome and Epidemiology Study (KoGES) showed that participants in the highest PRS quartile had a two-fold increased risk of obesity and HTN compared to those in the lowest quartile [31]. Similarly, using EWAS data of nearly 5,000 Europeans and Africans, a MRS constructed from 33 CpG loci accounted for 3.31% and 3.99% of the variance in SBP and DBP, respectively [32]. Additionally, a MRS based on 435 CpGs, derived from penalized regression of methylation data from 2,562 unrelated participants in Generation Scotland, explained around 10% of BMI variance, with each standard deviation increase in MRS associated with a 37% higher risk of obesity [33]. Furthermore, by contrasting MRS with PRS, a recent review emphasized the importance of integrating genetic and epigenetic data for improved trait prediction [29]. Indeed, studies combining PRS and MRS have demonstrated an increase in the explained variance in BMI, up to 14% [34] and 19% [13], highlighting the potential of multi-omics approaches.

Despite these achievements, as a comorbidity with various disorders, the prediction of OrH remains underexplored. The construction of PRS and MRS for OrH faces several challenges, especially in the Chinese population. One major limitation is the reduced efficacy of PRS and MRS when developed in one ancestry group and applied to others. To date, well-powered GWAS and EWAS have predominantly focused on individuals of European ancestry, limiting their applicability to other populations [10, 35–38]. Moreover, cultural and environmental factors unique to the Chinese population may influence how genetic variations and epigenetic modifications contribute to disease risk [10, 37].

On the other hand, multiple approaches have been developed for constructing PRS and MRS. In addition to the classic clumping and thresholding (C+T) method [39], shrinkage methods such as SCT [40], PRScs [41], ldpred2 [42, 43], and lassosum [44] adjust the weight of SNPs based on LD information. Other methods, such as PRScsx [45], CT-SLEB [46], PolyPredP^+^ [47], JointPRS [48], and PROSPER [49], are specifically designed for deployment across multiple ancestries, enhancing generalizability across diverse populations. For MRS construction, common computations approaches include C+T [34] and penalized linear regression [14]. Furthermore, integrating both PRS and MRS effectively could offer a promising opportunity to improve risk prediction for OrH.

In attempt to accurately assess OrH in the Chinese population, this study aimed to construct an integrative multi-omics model. Using data from 3,021 individuals in the Chinese Academy of Sciences (CAS), we evaluated various PRS methodologies based on GWAS statistics from the Biobank Japan (BBJ) and the UK Biobank (UKBB). Simultaneously, we analyzed several MRS models based on prior EWAS findings using data from 3,513 individuals in the National Survey of Physical Traits (NSPT). The performance of both PRS and MRS in predicting OrH risk was further validated in a separate dataset of 1,071 individuals from the CAS.

## Results

### Higher obesity, HTN, and OrH risk in male academics

Our study included a total of 7,605 Chinese individuals from two cohorts: 991 participants of Phase 1 and 3,101 of Phase 2 from the CAS cohort with phenotype and genomic data, and 3,513 participants from the NSPT cohort with phenotype and DNA methylation data. Additionally, methylation data were available for 1,071 samples in Phase 2 of the CAS cohort. According to the baseline data in **Table 1**, both the CAS and NSPT cohorts are middle-aged (average ages of 39.42±10.11 years and 50.21±12.75 years, respectively), with slightly fewer males (47.8% and 37.1%, respectively). A notable feature of the CAS cohort is the high proportion of participants with higher education (99.4% compared to 12.5% in the NSPT cohort).

**Table 1.**
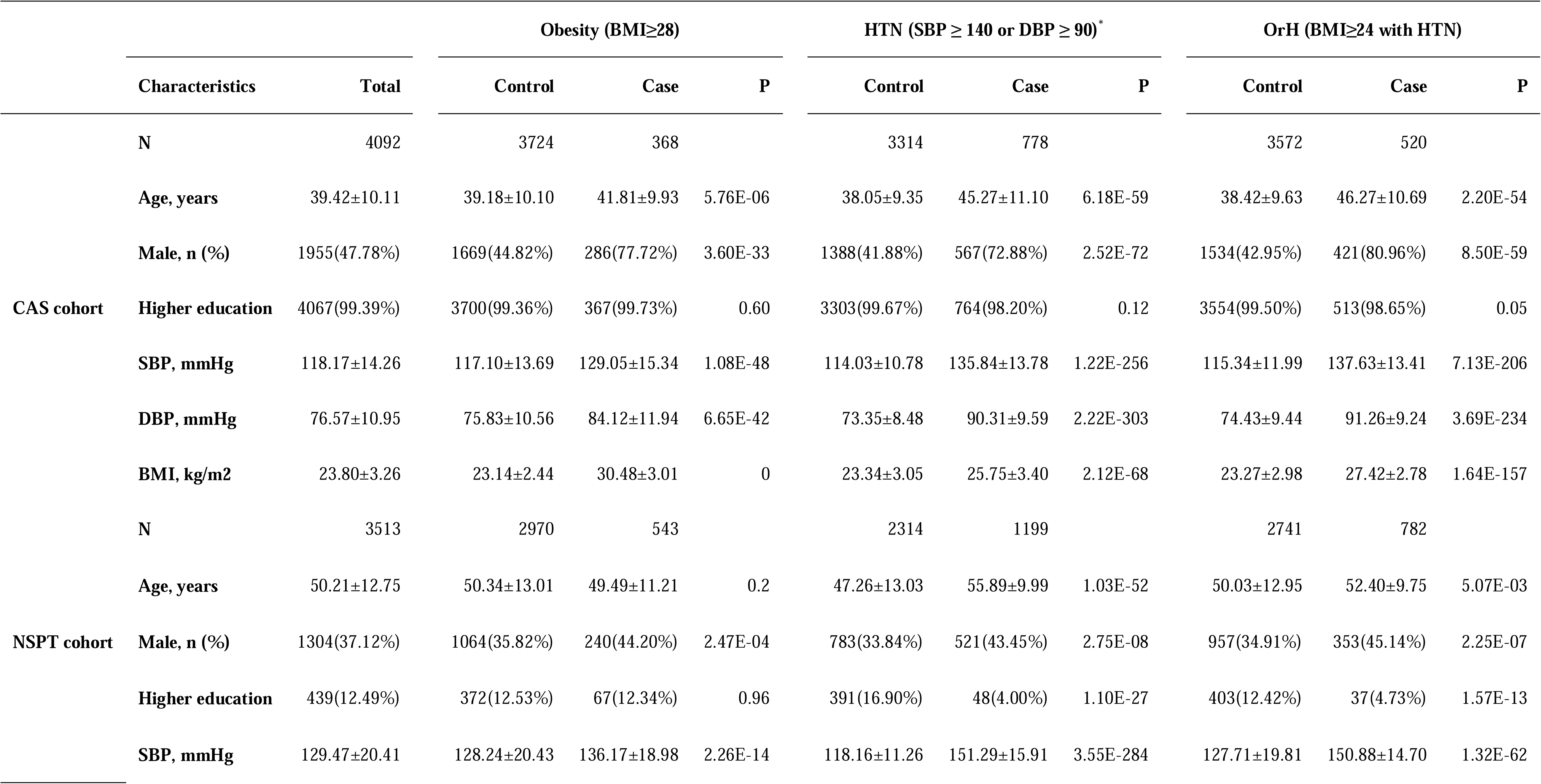
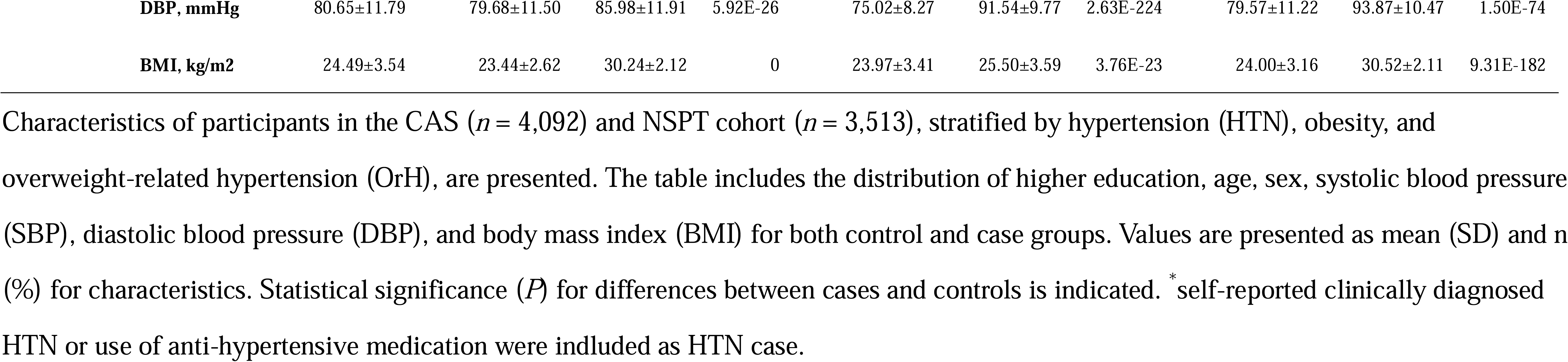
Baseline characteristics of participants in the study.

|  |  |  | Obesity (BMI≥28) |  |  | HTN (SBP ≥ 140 or DBP ≥ 90)* |  |  | OrH (BMI≥24 with HTN) |  |  |
| --- | --- | --- | --- | --- | --- | --- | --- | --- | --- | --- | --- |
|  | Characteristics | Total | Control | Case | P | Control | Case | P | Control | Case | P |
| CAS cohort | N | 4092 | 3724 | 368 |  | 3314 | 778 |  | 3572 | 520 |  |
|  | Age, years | 39.42±10.11 | 39.18±10.10 | 41.81±9.93 | 5.76E-06 | 38.05±9.35 | 45.27±11.10 | 6.18E-59 | 38.42±9.63 | 46.27±10.69 | 2.20E-54 |
|  | Male, n (%) | 1955(47.78%) | 1669(44.82%) | 286(77.72%) | 3.60E-33 | 1388(41.88%) | 567(72.88%) | 2.52E-72 | 1534(42.95%) | 421(80.96%) | 8.50E-59 |
|  | Higher education | 4067(99.39%) | 3700(99.36%) | 367(99.73%) | 0.60 | 3303(99.67%) | 764(98.20%) | 0.12 | 3554(99.50%) | 513(98.65%) | 0.05 |
|  | SBP, mmHg | 118.17±14.26 | 117.10±13.69 | 129.05±15.34 | 1.08E-48 | 114.03±10.78 | 135.84±13.78 | 1.22E-256 | 115.34±11.99 | 137.63±13.41 | 7.13E-206 |
|  | DBP, mmHg | 76.57±10.95 | 75.83±10.56 | 84.12±11.94 | 6.65E-42 | 73.35±8.48 | 90.31±9.59 | 2.22E-303 | 74.43±9.44 | 91.26±9.24 | 3.69E-234 |
|  | BMI, kg/m2 | 23.80±3.26 | 23.14±2.44 | 30.48±3.01 | 0 | 23.34±3.05 | 25.75±3.40 | 2.12E-68 | 23.27±2.98 | 27.42±2.78 | 1.64E-157 |
| NSPT cohort | N | 3513 | 2970 | 543 |  | 2314 | 1199 |  | 2741 | 782 |  |
|  | Age, years | 50.21±12.75 | 50.34±13.01 | 49.49±11.21 | 0.2 | 47.26±13.03 | 55.89±9.99 | 1.03E-52 | 50.03±12.95 | 52.40±9.75 | 5.07E-03 |
|  | Male, n (%) | 1304(37.12%) | 1064(35.82%) | 240(44.20%) | 2.47E-04 | 783(33.84%) | 521(43.45%) | 2.75E-08 | 957(34.91%) | 353(45.14%) | 2.25E-07 |
|  | Higher education | 439(12.49%) | 372(12.53%) | 67(12.34%) | 0.96 | 391(16.90%) | 48(4.00%) | 1.10E-27 | 403(12.42%) | 37(4.73%) | 1.57E-13 |
|  | SBP, mmHg | 129.47±20.41 | 128.24±20.43 | 136.17±18.98 | 2.26E-14 | 118.16±11.26 | 151.29±15.91 | 3.55E-284 | 127.71±19.81 | 150.88±14.70 | 1.32E-62 |

|  |  |  |  |  |  |  |  |  |  |  |
| --- | --- | --- | --- | --- | --- | --- | --- | --- | --- | --- |
| **DBP, mmHg** | 80.65±11.79 | 79.68±11.50 | 85.98±11.91 | 5.92E-26 | 75.02±8.27 | 91.54±9.77 | 2.63E-224 | 79.57±11.22 | 93.87±10.47 | 1.50E-74 |
| **BMI, kg/m2** | 24.49±3.54 | 23.44±2.62 | 30.24±2.12 | 0 | 23.97±3.41 | 25.50±3.59 | 3.76E-23 | 24.00±3.16 | 30.52±2.11 | 9.31E-182 |

Characteristics of participants in the CAS ( $n = 4,092$ ) and NSPT cohort ( $n = 3,513$ ), stratified by hypertension (HTN), obesity, and overweight-related hypertension (OrH), are presented. The table includes the distribution of higher education, age, sex, systolic blood pressure (SBP), diastolic blood pressure (DBP), and body mass index (BMI) for both control and case groups. Values are presented as mean (SD) and n (%) for characteristics. Statistical significance ( $P$ ) for differences between cases and controls is indicated. \* self-reported clinically diagnosed HTN or use of anti-hypertensive medication were included as HTN case.

A distinct pattern is observed in the CAS cohort, where obesity, HTN, and OrH exhibit a notably higher male-to-female (M/F) prevalence ratio compared to both the NSPT cohort and broader Chinese epidemiological studies. In CAS, as shown in **Table 1**, the proportion of males is 77.7% in obesity cases, 72.9% in HTN, and 81.0% in OrH. When converted to gender ratios, the male-to-female prevalence (M/F) ratio for obesity is 3.8 (14.6% vs. 3.8%, *P* = 3.60E-33), for HTN is 2.9 (29.0% vs. 9.9%, *P* = 2.52E-72), and the M/F ratio for OrH is particularly concerning at 4.7 (21.5% vs. 4.6%, *P* = 8.50E-59). Notably, this gender disparity persists across all age groups (**Figure 1**). In contrast, the proportions of males in obesity, HTN, and OrH cases are all lower in NSPT cohort (44.2%, 43.5%, and 45.1%, respectively), with the gender disparity (M/F ratio) being much less pronounced (1.3, 1.3 and 1.4, respectively). The gender ratio of NSPT is similar to that observed in the national survey on obesity and HTN (**Table S3**) [50–55].

**Figure 1.**
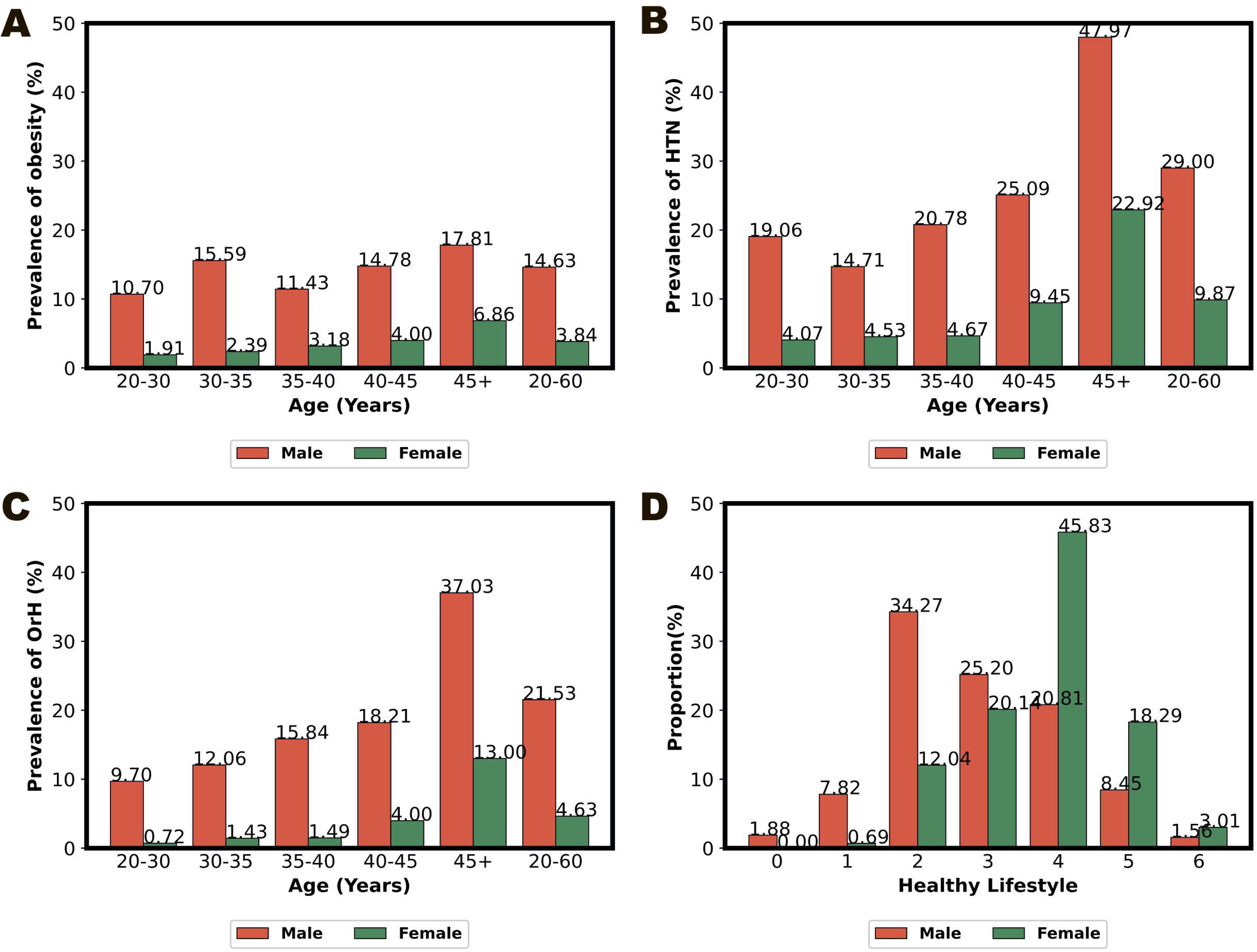
Prevalence of obesity, hypertension (HTN), and overweight-related hypertension (OrH) across age groups and healthy lifestyle scores by gender. Prevalence of obesity (A), HTN (B), and OrH (C) in males (red) and females (green) across different age groups in the CAS cohort (*n* = 4,092). (D) Distribution of healthy lifestyle scores by gender in the CAS1k multiomics cohort (*n* = 1,071). The healthy lifestyle score, ranging from 0 (least healthy) to 6 (most healthy), is calculated as the sum of six binary criteria: non-smoking, no excessive alcohol consumption, daily intake of fruits and vegetables, regular physical activity, BMI between 18.5 and 24, and waist circumference <85 cm for males and <80 cm for females.

Additionally, compared to NSPT cohort, gender disparities in metabolic health were more pronounced in CAS. CAS males exhibited significantly worse levels of multiple metabolic syndrome indicators, including total cholesterol (TC), triglycerides (TG), low-density lipoprotein (LDL), and high-density lipoprotein (HDL, **Table S4**). In contrast, CAS females demonstrated significantly better indicators, including TG, HDL, LDL, and fasting blood glucose (FBG), compared to NSPT females. These findings highlight a notable gender difference in the health conditions of academics in China.

Furthermore, we assessed healthy lifestyle scores in CAS cohort, calculated as the sum of six binary indicators (**Table S5**). As demonstrated in **Figure 1**, as high as 67.1% of women have their scores ≥4 compared to only 30.8% of men reach this score (*P* = 2.54E-31). Normal BMI (18.5–23.9 kg/m²) and waist circumference (WC, <85 cm for men and <80 cm for women) showed the most notable gender differences: BMI (34.4% in men vs. 66.2% in women, *P* = 3.12E-24) and WC (35.5% in men vs. 74.3% in women, *P* = 2.84E-35). Significant differences were also observed in the absence of current smoking (81.1% in men vs. 99.3% in women, *P* = 1.36E-19) and the absence of excessive drinking (92.6% in men vs. 98.8% in women, *P* = 7.30E-06).

### PRScsx outperforms other PRS methods

In this analysis, we utilized sub-datasets from the CAS cohort for PRS tuning, testing, and validation (see **Methods**), including: the PRS tuning set (*n* = 2,030, Phase 2 without CAS1k), PRS testing set (*n* = 991, Phase 1), and the validation set (*n* = 1,071, CAS1k). For quantitative traits including BMI, DBP, and SBP, 10 PRS methods (C+T, SCT, PRScs, ldpred2, lassosum, PRScsx, CT-SLEB, PolyPredP^+^, JointPRS and PROSPER) were trained in the PRS tuning set using GWAS summary statistics from the UKBB (*n* ≈ 450,000, European) and BBJ (*n* ≈ 150,000, Japanese, **Table S6**, **Figure S1** and **S2**). The latter five methods (PRScsx, CT-SLEB, PolyPredP^+^, JointPRS and PROSPER) represent multi-ancestry PRSs, where weights were derived by integrating UKBB and BBJ GWAS data.

Overall, throughout the tuning-testing-validation process (**Table S7-12**), the PRS generated using PRScsx method demonstrated robust performance, showing relatively strong prediction ability for the residuals of age-, sex-, and six genomic PC regressed quantitative phenotypes, including BMI, SBP, and DBP. Specifically, PRScsx achieved an R^2^ of 2.40%-9.81% in predicting residual variance, with an average of 4.76% in testing set (slightly lower than 4.87% for PROSPER) and 5.54% in validation set (higher than 4.98% for PROSPER, **Figure 2A, Table S12**).

**Figure 2.**
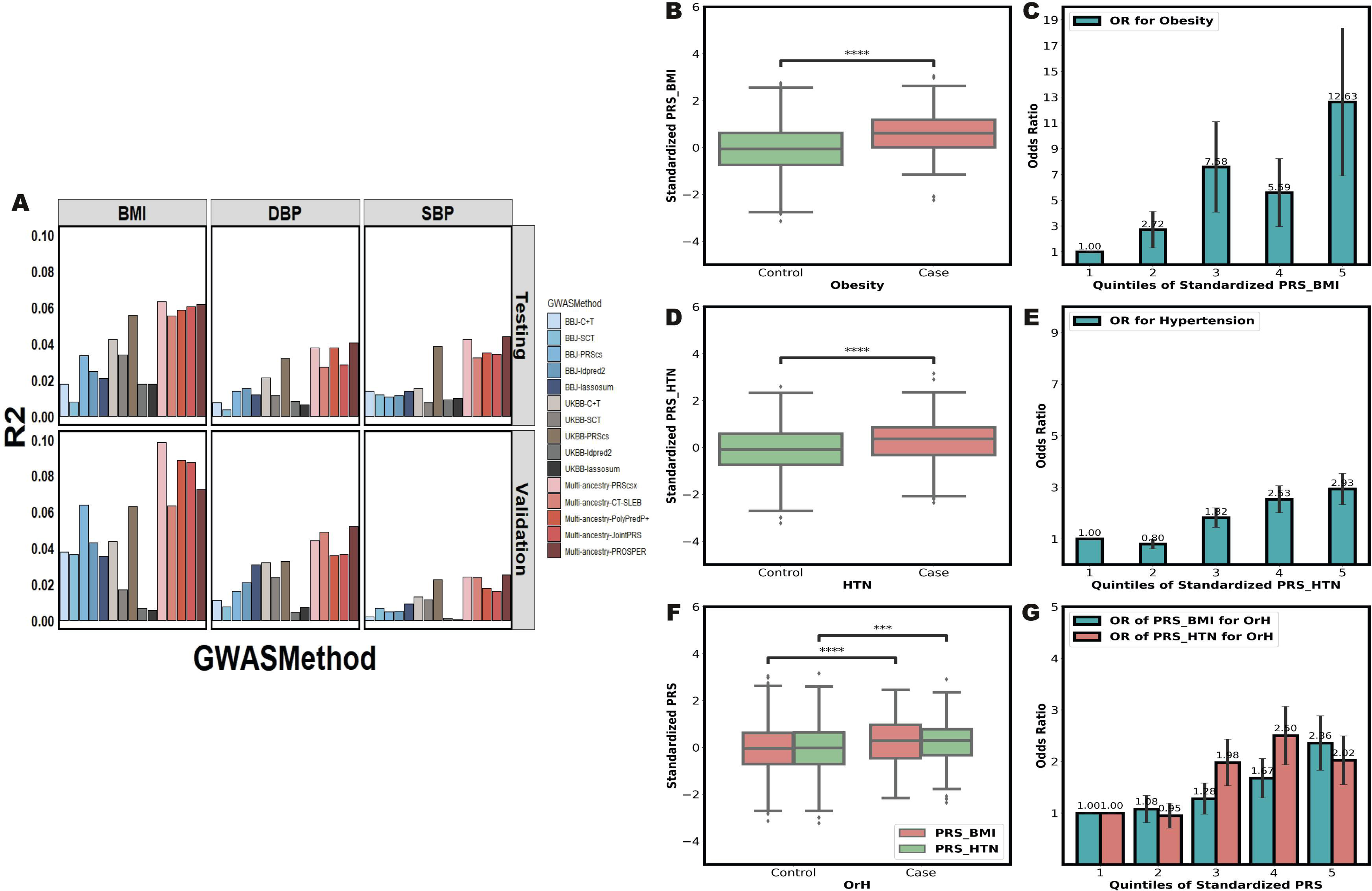
Performance of PRSs for BMI, DBP, and SBP as well as their association with obesity, HTN, and OrH. **(A)** The R² values (y-axis) represent the proportion of phenotypic variance explained by PRS for BMI, DBP, and SBP across different combinations of GWAS and methods (x-axis) in the testing dataset (*n* = 991, upper panel) and the validation dataset (*n* = 1,071, lower panel). Each bar corresponds to a specific method, as indicated by the color-coded legend. The phenotype was regressed on age, sex, and six PCs, and the residual from this regression was used as the dependent variable in the PRS modeling analyses. (**B)** Boxplot of PRS for BMI (PRS_BMI) in control vs. obesity case groups. (**C)** Odds ratios (OR) for obesity across quintiles of PRS_BMI, with the lowest 20% quintile serving as the reference group. (**D)** Boxplot of PRS for HTN (PRS_HTN) in control vs. obesity case groups. (**E)** OR for HTN across quintiles of PRS_HTN, with the lowest 20% quintile serving as the reference group. (**F)** Boxplot of PRS for BMI (PRS_BMI) and HTN (PRS_HTN) in control vs. OrH case groups. (**G)** OR for OrH across quintiles of PRS_BMI and PRS_HTN, with the lowest 20% quintile serving as the reference group. \*\*\*\**P* < 0.0001, \*\*\**P* < 0.001, \*\**P* < 0.01, \**P* < 0.05.

When compared to the PRSs from the PGS Catalog, the PRScsx method showed strong performance across multiple traits (BMI, SBP, and DBP) in both testing and validation sets (**Table S13**). The exceptions were DBP in the testing set, where PRScsx ranked second, slightly lower than that PGS003964 (3.75% vs. 4.90%), and SBP, where PRScsx ranked second in the testing set and third in the validation set, with marginal differences from the top-performing PRS. These results underscore the broad applicability and robustness of PRScsx in capturing the genetic architecture of complex traits, which led us to choose PRScsx for the subsequent analysis.

### Prediction accuracy of PRScsx for quantitative and binary traits

After comparison and selection of PRScsx, we further assessed its prediction performance in the validation dataset (CAS1k, *n* = 1,071) in predicting quantitative traits (BMI, SBP, and DBP) and binary disease outcomes (obesity, HTN, and OrH). The PRSs were largely normally distributed (Kolmogorov-Smirnov normality test with Bonferroni correction for multiple comparisons, *P* > 0.05).

For quantitative traits, our baseline prediction models, which incorporated sex and age only, revealed R² values of 18.28% for BMI, 17.86% for DBP, and 20.47% for SBP. When these models were augmented with respective PRScsx, there was a significant increase in accuracy, evidenced by R² values of 26.54% for BMI, 21.21% for DBP, and 22.71% for SBP (**Table 2**).

**Table 2.** The performance of different models for obesity, HTN and OrH in the validation set.

| Model | BMI | SBP | DBP | Obesity | HTN | OrH |
| --- | --- | --- | --- | --- | --- | --- |
| Baseline model | 18.28% [13.79%-22.48%] | 20.47% [16.02%-24.63%] | 17.86% [13.69%-21.84%] | 0.55[0.51-0.59] | 0.70[0.66-0.73] | 0.71[0.68-0.75] |
| PRS model | 26.54% [21.71%-31.01%] | 22.71% [18.10%-26.81%] | 21.21% [16.51%-25.31%] | 0.75[0.70-0.79] | 0.74[0.71-0.77] | 0.75[0.72-0.78] |
| MRS model | 26.86% [22.52%-31.00%] | 25.72% [20.87%-29.89%] | 23.15% [18.42%-27.42%] | 0.70[0.65-0.74] | 0.73[0.69-0.76] | 0.78[0.74-0.81] |
| Multi-omic model | 33.98% [29.12%-38.52%] | 27.58% [22.76%-31.61%] | 26.02% [21.42%-30.61%] | 0.77[0.72-0.81] | 0.76[0.73-0.79] | 0.80[0.77-0.83] |

The performance of baseline model (only with age and sex), PRS model (with age, sex and PRS), MRS model (with age, sex and MRS) and multi-omic model (with age, sex, PRS and MRS) was evaluated using R-squared ( $R^2$ ) for BMI, DBP and SBP and the AUC for obesity, HTN and OrH. All metrics and 95% confidence intervals were determined using a five-fold cross-validation approach in validation set ( $n = 1,071$ ).

Similarly, for binary disease statuses including obesity, HTN, and OrH, models integrated with PRScsx also demonstrated improved accuracy compared to baseline models. The PRS*_BMI_* distinctively segregated obesity from non-obesity groups (*P* = 4.35E-11, **Figure 2B**), with the OR for obesity increased by 2.13 for each standard deviation increment (OR/SD) in PRS*_BMI_* (95% CI: 1.77-2.57, *P* = 1.70E-11, **Table S15**). In a five-quantile schema, the OR for obesity rose progressively across quintiles, reaching 12.63 in the highest quintile (**Figure 2C**, **Table S15**). This inclusion of PRS*_BMI_* notably enhanced the model’s accuracy for obesity prediction, increasing the AUC from 0.55 to 0.75 (**Table 2**). For HTN, the combined PRS*_SBP_* and PRS*_DBP_* (PRS*_HTN_*) showed significant differentiation between HTN and non-HTN groups (*P* = 2.24E-08, **Figure 2D**), with an OR/SD of 1.68 in PRS*_HTN_* (95% CI: 1.47-1.91, *P* = 1.04E-10, **Table S15**). The ORs for HTN increased gradually across the five quantiles, peaking at 2.93 in the highest quintile (**Figure 2E**, **Table S15**). The inclusion of PRS*_HTN_* in the model resulted in an improvement in prediction accuracy for HTN (AUC increased from 0.70 to 0.74, **Table 2**).

Lastly, the model for predicting OrH showed a significant improvement in accuracy when including both PRS*_BMI_* and PRS*_HTN_* as predictors, with the AUC increasing from 0.71 to 0.75 (**Table 2**). Both of PRS*_BMI_* and PRS*_HTN_* demonstrated significant differentiation between OrH and non-OrH groups (*P* = 7.72E-05, *P* = 5.16E-04, **Figure 2F**), with an OR/SD of 1.42 in PRS*_BMI_* (95% CI: 1.23-1.63, *P* = 3.88E-5, **Table S15**) and 1.45 in PRS*_HTN_* (95% CI: 1.26-1.66, *P* = 1.63E-5, **Table S15**). In the highest quintile, the odds ratios for OrH peaked at 2.36 and 2.02 for PRS*_BMI_* and PRS*_HTN_*, respectively (**Figure 2G**, **Table S15**).

We further compared gender disparities in above analysis and no statistically significant differences were detected between males and females (T-test with Bonferroni correction for multiple comparisons, *P* > 0.05, **Figure S3A,B**), suggesting little genetic influence to the higher risk in men as observed above.

### Lasso MRS outperforms linear models and contributes to OrH risk profiling

Three MRS methods (Lasso, Linear 1, and Linear 2) were compared using distinct NSPT sub-dataset for MRS tuning (*n* = 2,047) and testing (*n* = 1,466), as well as the validation set for final examination (*n* = 1,071, see **Methods**). We focused on 1,506 CpGs for BMI, 77 for DBP, and 107 for SBP by reviewing prior EWAS results (**Table S11**) [13–25]. All MRSs were normally distributed (Kolmogorov-Smirnov normality test with Bonferroni correction for multiple comparisons, *P* > 0.05). Among the three methods, lasso achieved the best performance across all phenotypes in MRS testing set (R^2^_BMI_ = 8.48%, R^2^_DBP_ = 1.61%, R^2^_SBP_ = 3.02%,) and validation set (R^2^_BMI_ = 10.03%, R^2^_DBP_ = 4.68%, R^2^_SBP_ = 3.70%, **Figure 3A**, **Table S14**).

**Figure 3.**
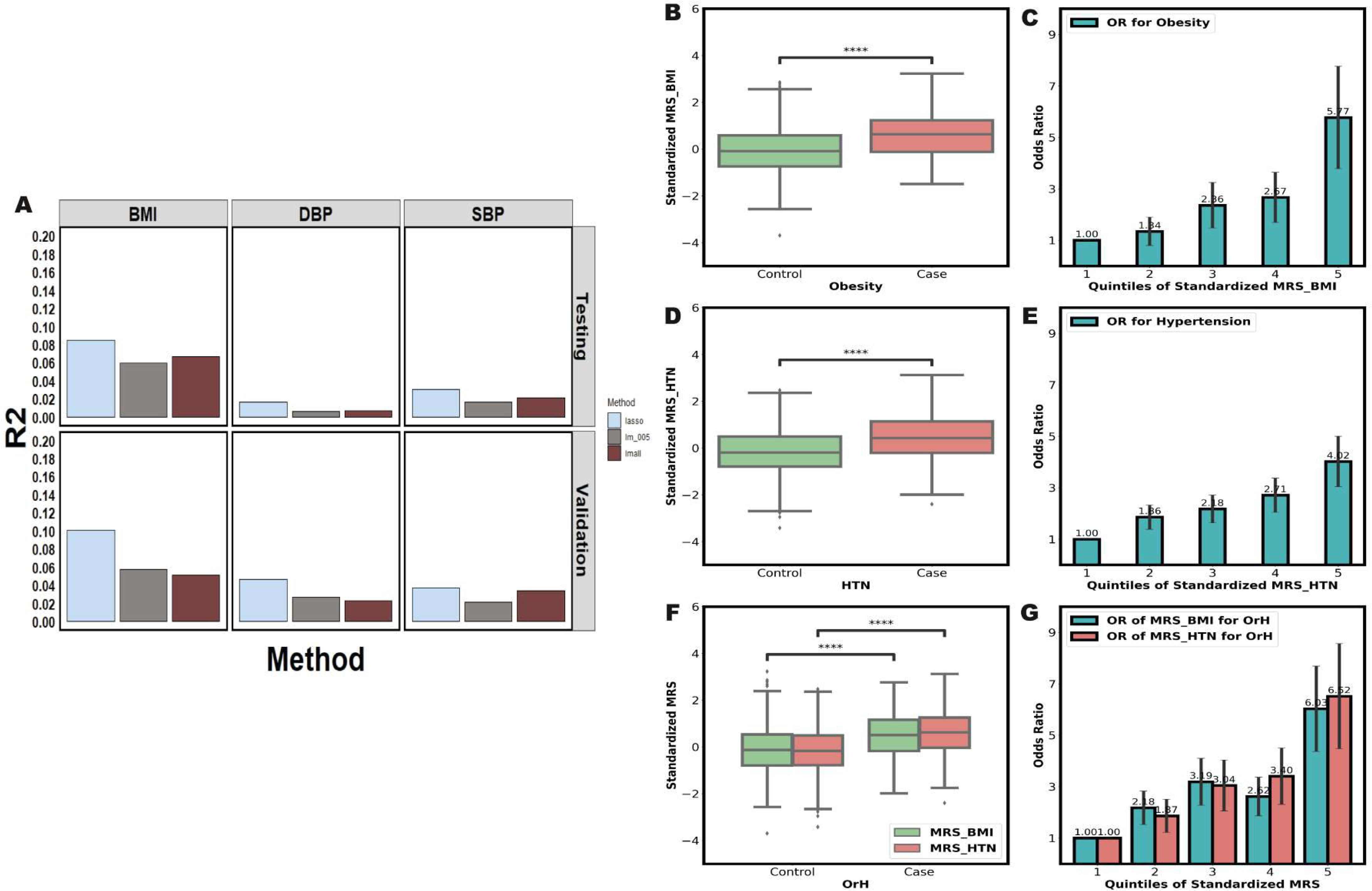
Performance of MRSs for BMI, DBP, and SBP as well as their association with obesity, HTN, and OrH. **(A)** The R² values (y-axis) represent the proportion of phenotypic variance explained by the MRSs for BMI, DBP, and SBP across different methods (x-axis). Results are shown for the testing dataset (*n* = 1,466, upper panel) and the validation dataset (*n* = 1,071, lower panel). Each bar corresponds to a specific method, as indicated by the color-coded legend. The phenotype was regressed on age, sex, and cell component proportions, and the residual from this regression was used as the dependent variable in the MRS modeling analyses. (**B)** Boxplot of MRS for BMI (MRS_BMI) in control vs. obesity case groups. (**C)** Odds ratios (OR) for obesity across quintiles of MRS_BMI, with the lowest 20% quintile serving as the reference group. (**D)** Boxplot of MRS for HTN (MRS_HTN) in control vs. obesity case groups. (**E)** OR for HTN across quintiles of MRS_HTN, with the lowest 20% quintile serving as the reference group. (**F)** Boxplot of MRS for BMI (MRS_BMI) and HTN (MRS_HTN) in control vs. OrH case groups. (**G)** OR for OrH across quintiles of MRS_BMI and MRS_HTN, with the lowest 20% quintile serving as the reference group. \*\*\*\**P* < 0.0001, \*\*\**P* < 0.001, \*\**P* < 0.01, \**P* < 0.05.

When compared to the baseline models that included sex and age as predictors, the lasso MRS showcased enhanced accuracy in validation set. Specifically, R² values increased from 18.28% to 26.86% for BMI, from 17.86% to 23.15% for DBP, and from 20.47% to 25.72% for SBP (**Table 2**). Notably, the MRS*_BMI_* exhibited a substantial difference between obesity and non-obesity individuals (*P* = 6.09E-11, **Figure 3B**) and an OR/SD of 1.88 for obesity (95% CI: 1.57-2.24, *P* = 4.94E-9, **Table S15**). In a five-quantile schema, the OR for obesity rose progressively across quintiles, reaching 5.77 in the highest quintile (**Figure 3C**, **Table S15**). This integration improved the AUC for obesity from 0.55 to 0.70 (**Table 2**). Similarly, MRS *_HTN_* revealed a clear differentiation between HTN and non-HTN groups (*P* = 6.87E-21, **Figure 3D**), with an OR/SD of 1.65 (95% CI: 143-1.89) (*P* = 3.85E-09, **Table S15**). The ORs for HTN increased gradually across the five quantiles, peaking at 4.02 in the highest quintile (**Figure 3E, Table S15**). This integration improved the AUC for HTN from 0.70 to 0.73 (**Table 2**). When considering both MRS*_BMI_* and MRS*_HTN_* in the OrH model, there was an increase in AUC from 0.71 to 0.78 (**Table 2**). Both of MRS*_BMI_* and MRS*_HTN_* demonstrated significant differentiation between OrH and non-OrH groups (*P* = 7.17E-16, *P* = 3.83E-23, **Figure 3F**), with an OR/SD of 1.52 in MRS*_BMI_* (95% CI: 1.31-1.77, *P* = 4.42E-06, **Table S15**) and 1.72 in MRS*_HTN_* (95% CI: 1.46-2.02, *P* = 4.99E-8, **Table S15**). In the highest quintile, the odds ratios for OrH peaked at 6.03 and 6.52 for MRS*_BMI_* and MRS*_HTN_*, respectively (**Figure 3G**, **Table S15**).

Notably, males exhibited significantly higher MRS values than females for BMI and blood pressure (5.08E-4 < *P* < 1.20E-41, **Figure S3**). These disparities may suggest the distinct influence of life styles and environmental exposures between genders in this cohort.

### Impact of metabolic and lifestyle factors on MRS predictions

We conducted a grouping analysis on the MRS-predicted values to assess whether other metabolic and lifestyle factors were associated with discrepancies between the MRS predictions and the observed values of BMI, DBP, and SBP (**Table S16**). The results indicated that discrepancies between the predicted and observed values are indeed associated with specific metabolic and lifestyle factors. For the MRS predictions underestimated BMI group, participants tended to have healthier lipid profiles (higher HDL, lower TG and LDL) and better lifestyle scores. Conversely, overestimation group was associated with less favorable lipid profiles, higher FBG, and poorer lifestyle scores. Similar patterns were observed for DBP and SBP, where overestimation by the MRS was linked to higher TC, LDL, FBG, and lower lifestyle scores. These results indicate that metabolic health and lifestyle behaviors may influence the accuracy of MRS predictions for these cardiovascular risk factors.

### Multi-omics model improved OrH risk profiling

We further integrated MRS and PRS in a multi-omics score and assessed its performance in predicting risk of obesity, HTN, and OrH in the validation set according to a five-fold cross-validation design. For obesity, adding MRS*_BMI_* to the PRS model improved the AUC from 0.75 to 0.77 (**Figure 4A**, **Table 2**). This multi-omics score fairly classified 0.47% of the population as high risk (predicted probability > 0.6), who indeed showed a high prevalence of 80.00% (**Figure 4D**); meanwhile, it effectively identified 59.48% of the population as having a low risk (predicted probability < 0.1), who in fact had a low prevalence of 4.71%. Consequently, this indicates that our model is informative for 59.95% of the population in obesity risk profiling. For HTN, adding MRS*_SBP_* and MRS*_DBP_* to the PRS model improved the AUC from 0.74 to 0.76 (**Figure 4B**, **Table 2**). And it is informative for 31.75% of the population in HTN risk profiling, effectively classifying the high risk (8.50% > 0.6 with a 71.43% prevalence) and low risk (23.25% < 0.1 with a 6.02% prevalence) groups (**Figure 4E**). In particular, for OrH adding MRS*_BMI_*, MRS*_SBP_*, and MRS*_DBP_* to PRS boosted AUC from 0.75 to 0.80 (**Figure 4C**, **Table 2**). This was informative for 43.89% of the population and effectively classified high risk (4.30% > 0.6 with a 63.04% prevalence) and low risk (39.59% < 0.1 with a 4.25% prevalence) groups (**Figure 4F**).

**Figure 4.**
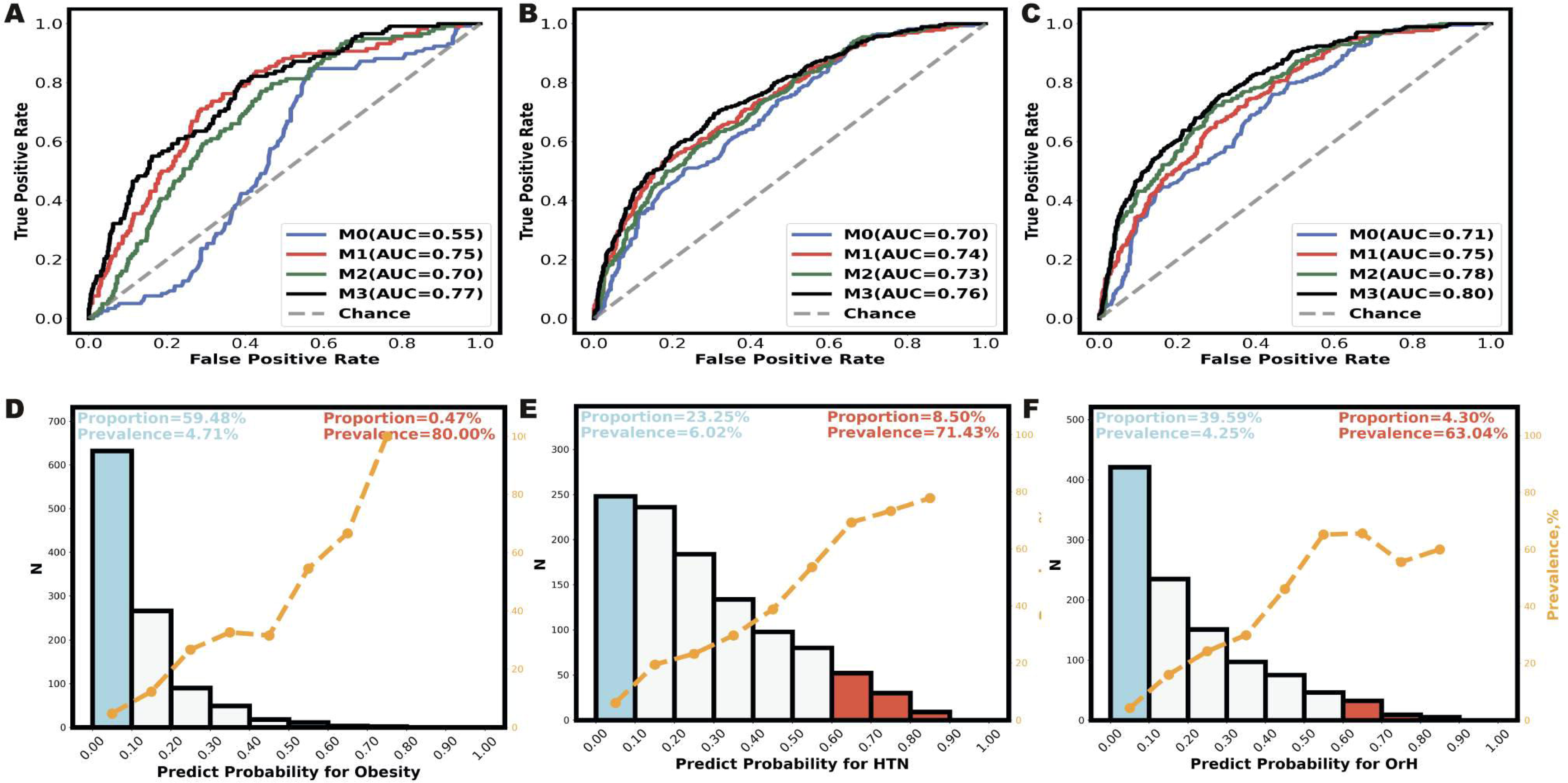
The prediction performance for obesity, HTN, and OrH using multi-omics models in the validation set. The Area Under the Receiver Operating Characteristic Curve (AUC) for obesity (**A**), HTN (**B**), and OrH (**C**) was assessed using a 5-fold cross-validated logistic regression model. The predictors included age and sex (M0), age, sex, and PRS (M1), age, sex, and MRS (M2), and age, sex, PRS, and MRS (M3) in the validation set (*n* = 1,071). In the multi-omics prediction model (M3), further focus was placed on individuals with extreme prediction probabilities for obesity (**D**), HTN (**E**), and OrH (**F**).The bars represent the number of participants within specific prediction probability intervals, with the blue bar indicating low risk (< 0.10) and the red bar indicating high risk (> 0.60). The orange line represents the prevalence in each interval.

## Discussion

In this study, we aimed to develop an accurate and effective prediction model for overweight-related hypertension by analyzing data from two general population cohorts, CAS and NSPT, totaling 7,605 individuals. We assessed the performance of ten methods for PRSs and three strategies for MRSs using a tuning-testing-validation approach. Additionally, we developed a multi-omics model to enhance prediction accuracy. Throughout our analysis, we also found distinct population characteristics among academics in our study.

### Factors related to gender disparity in disease prevalence among Chinese academics

One unexpected observation in our study was the notably high prevalence of obesity, HTN, and OrH among males in Chinese academics. Therefore we reviewed epidemiological data from the past three decades, which show significant changes in the prevalence of these conditions [50–55], most likely driven by China’s rapid economic growth and various risk factors, including economic status, gender, age, education, smoking, drinking, and inhabiting regions [56–61]. The male-to-female prevalence (M/F) ratio for obesity has fluctuated between 0.6 and 1.3 in both urban and rural areas, with a marked disparity in urban areas (M/F = 1.9) and even more pronounced among academics (M/F = 3.8, **Table S3**). For HTN, the M/F reached 2.9, significantly higher than in NSPT cohort and broader Chinese epidemiological studies, where it ranges from 1.1 to 1.3.

The gender disparity in disease prevalence observed among Chinese academics aligns with findings from several studies. Research from the China Health and Nutrition Survey and the Chinese National Center for Disease Control and Prevention indicated that women with higher education levels tend to have a lower BMI and reduced odds of being overweight, while men with higher education levels exhibit a higher BMI and increased odds of being overweight in China [62, 63]. Similar patterns have been observed in studies from BRICS economies [64] and Southern European countries [65, 66], further highlighting the association between education levels and obesity, particularly among women. Some studies have also examined the impact of education on HTN, indicating that individuals with higher education levels generally have healthier blood pressure. A Mendelian randomization study using data from FinnGen and the UK Biobank suggested a causal relationship between education level and HTN. For each standard deviation increase in genetically predicted higher education, the risk of HTN decreases by 44% [67]. Additionally, a study involving approximately 1.28 million adults from the China Health Evaluation and Risk Reduction through Nationwide Teamwork (ChinaHEART) project found that as education level increases, there is a significant downward trend in SBP [68].

To explore the potential reasons for the significant sex disparity, we first examined the genetic possibility and found no significant genetic differences in the performance of PRS. However, notable epigenetic differences were observed in MRS for the corresponding diseases between males and females. Consistent with previous studies linking higher MRS to poorer metabolic health [14, 36], our analysis showed that male academics exhibit higher MRS on BMI, DBP, and SBP, along with their relatively unhealthy lifestyles and metabolic syndrome traits. These findings suggest that the observed gender disparity is likely influenced by the combined effects of metabolic health and epigenetic factors.

We then briefly explored potential epigenetic explanations for the observed gender differences in disease prevalence. First, when examining regular physical activity, only minimal differences between sexes were found (**Table S5** ), which aligns with data of a 15-year national survey [69]. Thus, physical activity does not provide a compelling explanation for the gender disparities observed in the CAS cohort. However, for other lifestyle factors and metabolic syndrome profiles, male academics showed significantly poorer parameters compared to their female counterparts (**Table S4-5**). These differences are likely related to the more frequent social gatherings in males nationwide, such as dinners and drinking events, which are commonly associated with higher calories expenditure and alcohol consumption [70]. Additionally, men may experience great social pressure as the primary bread-winners for their families, which is also likely associated with unhealthy lifestyle choices and an increased risk of metabolic-related diseases [71–74]. Considering the dominant male composition in academia (especially in full professors), one possible reason for the notably high M/F ratio at CAS (3.8) could be the intense academic pressure and heavy workload in males, which may largely boost unhealthy lifestyle habits. On the other hand, cultural attitudes in China tend to favor slimmer figures for women [52], and female academics may demonstrate greater self-discipline regarding their health as well as more resources and opportunities to maintain their body shapes [60, 75], which further accentuate gender disparity in our cohort.

### Leveraging a multi-omics approach to enhance prediction analysis for OrH Profiling

OrH, as a common comorbidity pattern, would exacerbate cardiovascular and cerebrovascular damage more aggressively than simple obesity and HTN. Especially its prevalence has shown a significant upward trend globally in the past 20 years. Although the integration of PRS and MRS demonstrates improved utility in various diseases [76–78], there still exists a deficiency in multi-omics prediction models for OrH. In this study, we developed such an approach for OrH prediction using both genomic and epigenomic signals, achieving an AUC as high as 0.80.

We first developed effective PRSs for the Chinese population to predict BMI, DBP, and SBP by benchmarking five single-ancestry approaches (C+T, SCT, PRScs, LDpred2, and Lassosum) and five multi-ancestry approaches (PRScsx, CT-SLEB, PolyPredP^+^, JointPRS, and PROSPER). Among all methods, the top three with the highest accuracy are multi-ancestry methods across all traits. For BMI, the R² values for the top three multi-ancestry methods ranged from 8.74% to 9.81%, compared to the best single-ancestry model, which achieved 6.36% in validation analysis (**Table S12**). Similarly, for DBP the highest R² of multi-ancestry models was 5.18%, surpassing the best single-ancestry model of 3.28%. Our results well confirmed the outperformance of multi-ancestry PRS approaches over single-ancestry, and further demonstrated the enhanced generalizability of multi-ancestry PRS by leveraging shared genetic effects across different ancestries.

Among multi-ancestry PRS models, PRScsx consistently exhibited strong performance, achieving the highest R² for BMI and consistently ranking among the top three for both DBP and SBP in the validation set. This superior performance of complex traits across ancestries may be attributed to PRScsx’s advantage of Bayesian continuous shrinkage. Except for being only slightly (<0.4%) behind PGS003882 and PGS005015 for SBP in the validation set, the PRScsx model developed in our study notably outperforms many published models in the PGS Catalog (116 PRSs for BMI, 72 for SBP, and 51 for DBP). These results further emphasize the potential of our PRS profiling for the Chinese population as a robust and reliable tool for genetic risk prediction and precision medicine applications.

Unlike genetic models, methylation data provides a real-time snapshot of an individual’s risk profile by capturing the epigenetic landscape, which reflects not only genetic susceptibility but also modifiable influences that contribute to disease progression, as reported in numerous studies [79–82]. Therefore we aimed to use MRS for potential disease prediction based on currently available baseline data. After feature selection and optimization across various methylation models, lasso-MRS demonstrated the best performance in predicting BMI, DBP, and SBP. Our results yielded similar or better R^2^ compared to previous studies (e.g., 10.03% for BMI vs. 10.00% reported [33], and 4.68% for blood pressure vs. 3.99% [32]). Considering the environmental or lifestyle factors, these MRS models, especially with longitudinal data in the future, may provide valuable insights into an individual’s health status and potentially serve as early warnings for unhealthy conditions.

Moreover, combining MRS with PRS enhances risk prediction by linking genetic susceptibility with current epigenetic states. Indeed with the AUC of 0.75 for PRS and 0.78 for MRS, we observed an integrated AUC of 0.80 for OrH risk profiling, further confirming a shared molecular mechanism in obesity and hypertension. This profiling may also fill a critical gap in individualized early warning for cardiovascular and metabolic disorders. By identifying high-risk individuals (risk score > 0.6) using multi-omics models, such as the 4.30% for OrH in CAS cohort, healthcare providers can implement more targeted preventive measures and treatment strategies to improve their health status.

## Conclusion

This research reveals a notably high prevalence of obesity, HTN, and OrH among males but significantly lower prevalence among females in Chinese academics with characterizations of research career and higher education. These results considerably diverge from common patterns observed in Chinese epidemiological investigations. Additional analysis indicates such large gender disparities are primarily associated to the complex interplay among epigenetic factors, lifestyle and metabolic health, raising concerns about notably higher risks for males within Chinese academics. In omics analysis, PRScsx and lasso in MRS method demonstrates high potential as a robust tool for risk assessment of obesity, HTN, and OrH. The integration of PRS and MRS further enhance the accuracy of the risk profiling, suggesting the effectiveness of multi-omics approach for improved personalized risk assessment strategies especially for OrH high-risk populations.

## Materials and methods

### Study population – CAS cohort

This study involved 4,092 Chinese participants from the CAS cohort, which was established in 2015 to target employees of the Chinese Academy of Sciences (CAS) in the Beijing region. Informed consent was obtained from all participants, and the study protocol was approved by the Ethics Committee of the Beijing Institute of Genomics and associated hospitals. The cohort was highly educated, with 99.3% holding at least a university degree. Before undergoing clinical assessments, participants completed an online questionnaire that gathered information on factors such as gender, smoking status, alcohol consumption, tea intake, and sleep duration. Clinical health assessments, including anthropometric, physical, blood, urine, and imaging exams, were performed at designated hospitals, where 8 ml of blood was collected from each participant. The research protocol received approval from the Ethics Committee of the Beijing Institute of Genomics, Chinese Academy of Sciences (2015H023 and 2021H001), the Ethics Committee of Beijing Zhongguancun Hospital (20201229).

Recruitment occurred in two phases. Phase 1 (2015-2016) included 991 participants, whose DNA samples were analyzed using 30X whole-genome sequencing (WGS), and all phenotypic data were collected at the General Hospital of Aviation Industry Corporation of China. Phase 2 (2020-2021) added 3,101 participants, whose DNA samples were analyzed using Illumina genotyping microarrays, and phenotypic data were collected at Beijing Zhongguancun Hospital. In Phase 2, 1,071 individuals were designated as the CAS1k subgroup, designed to provide multi-omics data, with their samples analyzed using Illumina methylation microarrays.

### Study population – NSPT cohort

The National Survey of Physical Traits cohort (NSPT) is a population-based prospective cohort study consisted of 3,523 Han Chinese individuals from various regions of China, including Taizhou, Nanning, and Zhengzhou (1,310 males and 2,213 females, aged from 18 to 83 years old, mean ±s.d. = 50.21 ± 12.75). After quality control, 3,513 participants remained for analysis, which included three phases: Phase 1 (*n* = 690) in 2018, Phase 2 (*n* = 776) in 2019, and Phase 3 (*n* = 2,047) in 2019. DNA methylation was assessed using the Illumina methylation microarray on blood samples. The study was approved by the Ethics Committee of Shanghai Institutes for Biological Sciences (ER-SIBS-261410), and written informed consent was obtained from all participants.

### Definitions of overweight, obesity, HTN, OrH, healthy lifestyle and higher education

Overweight was defined as a BMI between 24.0 and 27.9 kg/m², while obesity was defined as BMI ≥ 28.0 kg/m² according to China’s guidelines [83]. HTN was defined as either SBP ≥ 140 mmHg, DBP ≥ 90 mmHg, self-reported HTN diagnosis, or use of antihypertensive medications. Individuals with both a BMI ≥ 24 and HTN were categorized as having OrH [84]. It is important to note that thresholds for defining obesity or HTN may vary across populations [85, 86], and comparisons with studies using different criteria should be interpreted with caution.

Healthy lifestyle factors were defined based on the China Kadoorie Biobank (CKB) criteria [87], which include not smoking, not engaging in excessive alcohol consumption, maintaining a healthy diet (daily fruit and vegetable intake), participating in regular physical activity, and having a BMI between 18.5 and 23.9 kg/m² and a waist circumference of <85 cm for males and <80 cm for females. Participants earned a score of 1 for each criterion they met and 0 for each one they did not, resulting in a total score ranging from 0 to 6, representing their overall healthy lifestyle. Higher education was defined as having any college or university degree.

### Whole genome sequencing and microarray genotyping in CAS Cohort

WGS was performed at 30X coverage using the Illumina HiSeq 3000 (Illumina, San, Diego, CA), and sequencing reads were aligned to the hg19 reference genome [88]. Variants were called using GATK [89] and annotated using ANNOVAR [90], with detailed methods for sample and library preparation reported previously [91].

Microarray genotyping was conducted using the Infinium Asian Screening Array + MultiDisease-24 Bead Chip. SNP genotypes were phased and imputed using the IMPUTE2 [92] based on the East Asian population in the 1000 Genomes Project [93, 94].

Quality control measures included removing individuals with gender mismatches, low genotyping call rates (<97%), or abnormal heterozygosity (outside the mean ± 3 SD range). For SNPs, we excluded those with imputation scores <0.6 (in the CAS Phase 2 cohort), Hardy-Weinberg equilibrium *P* < 1E-4, genotyping call rates <98%, and minor allele frequency (MAF) <1%. After these steps, 3,169,262 SNPs and 4,092 individuals were retained for analysis.

### Methylation microarray of CAS1k and NSPT cohort

Both the CAS1k and NSPT cohorts’ DNA methylation data were generated using the Illumina Infinium Methylation EPIC BeadChip. The raw array data were processed using the ChAMP package [95] in R to compute β values for methylation levels. Probes were filtered based on Illumina quality thresholds (bead count < 3 in >5% of samples and 1% of samples with a detection p value > 0.05). Batch effects were corrected using the ComBat [96, 97] method, and cell-type heterogeneity was adjusted using the EpiDISH method [98]. After quality control, 751,015 CpGs were retained for the CAS1k cohort, and 811,876 CpGs were retained for the NSPT cohort.

### Construction and selection of PRS

The PRS construction and selection followed a tuning-testing-validation design using the CAS cohort. The tuning set included 2,030 participants from Phase 2 (excluding CAS1k), the testing set had 991 participants from Phase 1, and the validation set consisted of 1,071 participants from CAS1k.

PRSs for BMI, DBP, and SBP were derived using GWAS summary statistics from the UKBB [99] (∼450,000 Europeans) and BBJ [100, 101] (∼150,000 Japanese). Ten PRS methods were applied: C+T [39], SCT [40], PRScs [41], ldpred2 [42, 43], lassosum [44], PRScsx [45], CT-SLEB [46], PolyPredP^+^ [47], JointPRS [48] and PROSPER [49]. Hyperparameters were fine-tuned in the tuning set and evaluated in the testing and validation sets, with performance assessed by R-squared and 95% confidence intervals using bootstrap resampling (k = 10,000, detailed in **Supplemental Methods**).

To assess the optimal PRS for East Asians, we compared it to existing scores in the PGS Catalog, selecting 48, 4, and 20 PRSs for BMI, DBP, and SBP, respectively, based on the required information. The best-performing PRS was then used for multi-omics prediction analysis in the validation set. In all PRS analyses, phenotypes were regressed on age, sex, and six genomic principal components, with residuals used for PRS modeling to calculate adjusted R² values reflecting variance explained beyond potential confounders.

### Construction and selection of MRS

The MRS construction and selection followed a tuning-testing-validation design using both NSPT and CAS1k cohort. The MRS tuning set included 2,047 participants from Phase 2 of the NSPT cohort, while the MRS testing set consisted of 1,466 participants from the Phase 1 and Phase 3. The validation set was made up of 1,071 individuals from CAS1k. MRSs for BMI, DBP, and SBP were derived from findings in previous studies [13–25] resulting in final sets of 1,506, 77, and 107 CpGs for BMI, DBP, and SBP, respectively (details provided in **Supplemental Methods**).

To construct the MRSs, we used three different methods: Linear Regression 1, which included all CpGs from the studies without filtering; Linear Regression 2, which selected only CpGs with a P-value < 0.05; and Lasso Regression, which applied penalized linear regression to optimize the model. Different CpG sets with corresponding beta coefficients were generated in the tuning set, then evaluated and validated in the testing and validation sets.

The best-performing MRS was selected and applied to the validation set for multi-omics prediction analysis. In all MRS analyses, phenotypes were regressed on age, sex, and cell composition, with the residuals serving as the dependent variable for MRS modeling. This approach enabled us to report adjusted R² values and 95% CI, calculated through bootstrap resampling (k = 10,000), that account for the variance in MRS explained beyond the confounding effects of age, sex, and cell composition.

### Grouping analysis on the MRS-predicted values

A grouping analysis on the MRS-predicted values was performed in validation set to determine whether discrepancies between the MRS predictions and observed values for BMI, DBP, and SBP could be associated with metabolic and lifestyle factors. Participants were grouped based on quantiles of the prediction error (predicted minus observed). The groups included the lowest 10% quantile, where the MRS significantly underestimated the trait; the middle 80% quantiles, where the MRS predictions closely matched the observed values; and the highest 10% quantile, where the MRS significantly overestimated the trait. Metabolic factors, including HDL, LDL, TC, TG, FBG, and healthy lifestyle score, were compared across these groups using independent t-tests to identify significant differences between the underestimated, accurately predicted, and overestimated groups.

### Multi-omics risk prediction

The best-performing PRS and MRS were combined into multi-omics scores, and their prediction performance was assessed in the validation set using linear regression for continuous traits (BMI, SBP, and DBP) and logistic regression for binary traits (obesity, HTN, and OrH). These models were adjusted for age and sex as covariates. The performance was evaluated using a five-fold cross-validation design, which was trained on four parts and tested on the remaining one, and this process is repeated five times, each time using a different part of the data for testing. The AUC are averaged across the five iterations to provide a more generalized estimate of the model’s prediction power. This approach ensured a consistent and robust comparison of baseline (only sex and age), PRS, MRS, and multi-omics approaches.

For continuous traits, separate models were developed using the corresponding PRS, MRS, or both (multi-omics) as predictors. For binary traits, obesity models used PRS*_BMI_*, MRS*_BMI_*, or both as predictors. HTN models incorporated either the average of PRS*_DBP_* and PRS*_SBP_*, the average of MRS*_DBP_* and MRS*_SBP,_* or both averages as predictors. For OrH, models included PRS*_BMI_* and the average of PRS*_DBP_* and PRS*_SBP_*, MRS*_BMI_* and the average of MRS*_DBP_* and MRS*_SBP_*, or all four predictors.

Model performance was assessed using R² for continuous traits and AUC for binary traits, calculated through 5-fold cross-validation within the validation set. All analyses were performed using R (version 4.0.3) and Python (version 3.6.4).

## CRediT author statement

**Yaning Zhang**: Conceptualization, Methodology, Formal analysis, Writing-Original draft preparation. **Qiwen Zheng**: Conceptualization, Visualization, Writing - Review & Editing. **Qili Qian**: Data Curation, Formal analysis. **Na Yuan:** Data Curation. **Tianzi Liu:** Data Curation. **Xingjian Gao:** Data Curation. **Xiu Fan:** Data Curation. **Youkun Bi:** Data Curation. **Guangju Ji:** Data Curation. **Peilin Jia:** Data Curation. **Sijia Wang**, **Fan Liu** and **Changqing Zeng**: Conceptualization, Supervision, Writing - Review & Editing. All authors have read and approved the final manuscript.

## Supporting information

supplementary matrial

supplementary matrial

## Acknowledgments

The authors thank the participants who contributed their data in the CAS cohort and the NSPT cohort study. This work was supported by the Science and Technology Service Network Initiative of Chinese Academy of Sciences (Grant number KFJ-STS-ZDTP-079), Strategic Priority Research Program of Chinese Academy of Sciences (Grant number XDB38010400), the Startup Research Fund of Henan Academy of Sciences (Grant number 232016009), the Scientific and Technological Research Project of Henan Province(Grant number 232102310066), the Basic research fund of Henan Academy of Sciences (Grant number 230618032), the Startup Research Fund of Henan Academy of Sciences (Grant number 231816040), the National Natural Science Foundation of China (NSFC) (Grant number 32325013), the CAS Project for Young Scientists in Basic Research (Grant number YSBR-077), the Strategic Priority Research Program of the Chinese Academy of Sciences (Grant number XDB38020400 to SW), and Shanghai Science and Technology Commission Excellent Academic Leaders Program (Grant number 22XD1424700).

## Data availability

Biobank Japan (BBJ) GWAS summary statistics [102] are publicly available on the BBJ website (https://pheweb.jp). UK Biobank (UKBB) GWAS summary statistics [99] are publicly available on the Gene ATLAS website (http://geneatlas.roslin.ed.ac.uk/). The CAS cohort and NSPT cohort phenotypic, genetic data and DNA methylation data that support the findings of this study are available from the corresponding author upon reasonable request.

## Ethics approval and consent to participate

The research protocol received approval from the Ethics Committee of the Beijing Institute of Genomics, Chinese Academy of Sciences (2015H023 and 2021H001), the Ethics Committee of Beijing Zhongguancun Hospital (20201229), the Ethics Committees of Fudan University (14117) and the Shanghai Institutes for Biological Sciences (ER-SIBS-261410). Participants provided written informed consent allowing use of their samples and data for medical research purposes, and these ethical regulations cover the work in this study. Written informed consent was obtained from all of the participants.

## Declaration of Generative AI and AI-assisted technologies in the writing process

During the preparation of this work the authors used GPT-4 in order to correct the English in the manuscript. After using this tool, the authors reviewed and edited the content as needed and take full responsibility for the content of the publication.

