## supplementary matrial for "An Integrative Polygenic and Epigenetic Risk Score for Overweight-Related Hypertension in Chinese Population"

**Title**

**Author(s) Address Information**

^1^ *Henan Academy of Sciences, Zhengzhou 450006, China*

^2^ *CAS Key Laboratory of Genomic and Precision Medicine, Beijing Institute of Genomics (National Center for Bioinformation), University of Chinese Academy of Sciences, Chinese Academy of Sciences, Beijing 100101, China*

^3^ *Department of Forensic Sciences, College of Criminal Justice, Naif Arab University for Security Sciences, Riyadh, Saudi Arabia*

^4^ *CAS Key Laboratory of Computational Biology, Shanghai Institute of Nutrition and Health, Chinese Academy of Sciences, Shanghai 200031, China*

^5^ *National Clinical Research Center of Kidney Diseases, Jinling Hospital, Nanjing, Jiangsu, China*

^6^ *Institute of Biophysics, Chinese Academy of Sciences, Beijing 100101, China*

^#^ Yaning Zhang and Qiwen Zheng and Qili Qian are the co-first authors of this article

### Supplementary Methods

#### UK Biobank and Biobank Japan genome-wide association study summary statistics

In order to generate different ancestry PRSs, we collected GWAS summary statistics through previous studies of East Asian and European separately to be associated with BMI, DBP and SBP. European GWAS data was collected from UK biobank (UKBB)[1] while East Asian GWAS data was collected from Biobank Japan (BBJ)[2, 3]. UKBB is a large-scale prospective cohort composed of about 500,000 of mainly European ancestry adults, whereas BBJ is a hospital-based disease-ascertained cohort with approximately 200,000 participants of mainly Japanese ancestry[4-6].

For the UK Biobank (UKBB) GWAS summary, we included only autosomal SNPs with a Minor Allele Frequency (MAF) greater than 1% and an INFO score exceeding 0.9, resulting in approximately 12 million SNPs for analysis. Similarly, for the BioBank Japan (BBJ) GWAS summary, we considered autosomal SNPs with MAF > 1% and Rsq > 0.9, yielding around 5.9 million SNPs for analysis. To ensure consistency in SNP magnitude across datasets, we performed analyses on the intersection of UKBB GWAS summary, BBJ GWAS summary, and the CAS cohort. Following the correction of positive and negative strands in the different datasets, we identified 3,169,089 autosomal markers available for subsequent analyses. To facilitate uniformity in GWAS summary statistics, we reformatted the data to include SNP ID, chromosome, base pair position, Effect Allele (EA), Non-Effect Allele (Non-EA), EA frequency, effect size estimate, effect size estimate standard error, and *P* value.

#### Construction and selection of polygenic risk scores

To generate various polygenic risk scores (PRS), we employed a total of 15 strategies using GWAS data from the UKBB and BBJ for BMI, DBP, and SBP. The construction of PRS involved utilizing GWAS summary statistics from both the UKBB or BBJ datasets. We employed 10 PRS construction methods, namely C+T[7], SCT[8], PRScs[9], ldpred2[10, 11], lassosum[12], PRScsx[13], CT-SLEB[14], PolyPredP+[15], JointPRS[16] and PROSPER[17]. Hyperparameters for each method were fine-tuned within the PRS tuning set to determine the optimal PRS for each trait.

For single-ancestry PRSs, five methods (C+T, SCT, PRScs, ldpred2, lassosum) were applied to either UKBB or BBJ data, resulting in a total of 10 PRSs. For the optimization for C+T, we use the large grid of hyper-parameters testing a threshold of clumping r^2^c within [0.01, 0.05, 0.1, 0.2, 0.5, 0.8, 0.95], a base size of clumping window within [50, 100, 200, 500] in kb where the actual window size is then computed as the base size divided by r^2^c, and a sequence of 20 thresholds on -log10(P-values) between 0.1 and the most significant P-value, equally spaced on a log scale. SCTPRS employed penalized regression to find the optimal linear combination of all C+T PRS. PRScs utilized a full Bayesian approach (a=1, b=0.5) to estimate the global shrinkage parameter, phi. For ldpred2, we tested 12 combinations of parameters, with p values selected from [0.003,0.01,0.03,0.1,0.3,1], h² estimated from LDSC, and the sparse option set to [FALSE,TRUE]. For lassosum, we explored 80 parameter combinations using a large grid of hyperparameters, testing delta thresholds from [0.001,0.01,0.1,1], with nlambda set to 20 and lambda.min.ratio fixed at 0.01.

In the case of multi-ancestry PRSs, GWAS data from both UKBB and BBJ were combined, and the PRScsx, CT-SLEB, PolyPredP+, JointPRS and PROSPER methods were applied, yielding five additional PRSs. Specifically, the PRS for PRScsx method for each phenotype integrated EAS and EUR GWAS summary statistics using the parameter, phi. a=1, b=0.5. For JointPRS, we used a linear combination of the PRSs with parameter (phi=auto, a=1, b=0.5) using EAS and EUR posterior SNP effect size estimates, taking the genetic correlation into consideration in addition. For CT-SLEB, we selected the best PRS from 2268 combinations using GWAS summary statistics from EAS and EUR, integrating clumping and thresholding, empirical Bayes and 3 super-learning models: glmnet, ridge, nnet. For PolyPredP+, we combinated of PolyFun-pred with PRScs-UKBB and PRScs-BBJ. And for PROSPER, PRS was developed using an ensemble of 50 combinations of delta, λ and c and 3 super-learning models: glmnet,SL.ridge and lm.

A reference panel of 503 European individuals from the 1000 Genome Project was used for all methods when deriving effect sizes from the UKBB GWAS. Additionally, 504 East Asian individuals from the 1000 Genome Project served as the LD reference for all methods when effect sizes were derived from the BBJ GWAS.

The optimized PRSs underwent evaluation in the PRS testing and validation set, assessing their performance through model fitness R^2^ and calculating the 95% confidence interval (CI) using bootstrap resampling (k=10,000). In all PRS modeling testing procedures, the phenotype was regressed on age, sex, and six genomic principal components. The residuals from this regression were then used as the dependent variable in PRS modeling and testing analyses.

#### CpG selection for MRS

Numerous CpG sites have been associated with BMI due to the multifaceted nature of metabolic syndrome, which includes hyperlipidaemia, hyperglycaemia, and inflammatory changes reflected in DNAm changes as highlighted by Bell et al. [18] and Do et al. [19]. We developed comprehensive lists of CpG sites for BMI, DBP, and SBP by systematically integrating findings from previous studies and applying rigorous quality control measures.

For BMI, we included 1,109 CpGs identified by McCartney et al. [20], retaining 1,075 after quality control. We further incorporated 397 CpGs reported by Do et al. [19], with 368 passing quality checks, along with 3 CpG sites discovered by Li et al. [21]. 2 CpGs identified by Dick et al. [22], 223 of 239 CpGs identified by Wahlet al. [23], and 2 CpGs from Chen et al. [24] were also included. After excluding overlapping CpGs across studies, the final list for BMI comprised 1,506 unique CpGs.

For DBP, we included 42 of 47 CpGs from Richard et al. [25], 26 of 28 CpGs from Kato et al. [26], and 19 of 20 CpGs from Hong et al. [27]. Following the removal of overlapping CpGs, the finalized list contained 77 unique CpGs for DBP.

For SBP, we incorporated 26 of 28 CpGs from Hoyt et al. [28], 1 CpG from Huan et al.[29], 2 CpGs from Si et al. [30], 16 of 17 CpGs from Hong et al. [27], 1 CpG from Kou et al. [31], and 73 of 77 CpGs from Richard et al. [25]. After accounting for overlaps, the final list included 107 unique CpGs for SBP.

### Supplemental Tables

#### Table S1 Summary of BMI, SBP and DBP associated SNPs from GWAS Catalog

Please refer to the MS Excel file entitled Supplementary Information.xlsx.

#### Table S2 Summary of BMI, SBP and DBP associated CpGs from published studies

Please refer to the MS Excel file entitled Supplementary Information.xlsx.

#### Table S3 Summary of research on the prevalence of HTN and obesity in Chinese adults

|  | **N** | **Adults prevalence** | **Male adults prevalence** | **Female adults prevalence** | **Male/Female** | **Year of investigation** | **Region** | **Data source** | **Reference PMID** |
| --- | --- | --- | --- | --- | --- | --- | --- | --- | --- |
| Obesity | 7303 | 4.2% | 3.0% | 5.2% | 0.6 | 1993 | Urban and rural | China Health and Nutrition Survey(CHNS) | 33037330 |
|  | 7716 | 6.5% | 5.7% | 7.3% | 0.8 | 1997 | Urban and rural |  |  |
|  | 8266 | 8.0% | 7.0% | 8.9% | 0.8 | 2000 | Urban and rural |  |  |
|  | 8656 | 9.2% | 8.4% | 9.9% | 0.8 | 2004 | Urban and rural |  |  |
|  | 8490 | 9.4% | 9.1% | 9.7% | 0.9 | 2006 | Urban and rural |  |  |
|  | 9051 | 10.7% | 10.8% | 10.6% | 1.0 | 2009 | Urban and rural |  |  |
|  | 11919 | 13.2% | 13.6% | 13.0% | 1.0 | 2011 | Urban and rural |  |  |
|  | 8841 | 15.7% | 17.9% | 14.4% | 1.2 | 2015 | Urban and rural |  |  |
|  | 19477 | 16.4% | 18.2% | 14.7% | 1.2 | 2018 | Urban and rural | The China Chronic Disease and Nutrition Surveillance (CCDNS) surveys | 34022156 |
|  | 1577094 | 14.1% | 18.2% | 9.4% | 1.9 | 2019 | Urban | Meinian Institute of Health | 37589256 |
|  | 3513 | 15.50% | 18.40% | 13.70% | 1.3 | 2015-2019 | Urban and rural | NSPT cohort | / |
|  | 4092 | 9.0% | 14.6% | 3.8% | 3.8 | 2015-2021 | Urban,  high educational professional | CAS cohort | / |
| HTN | 8567 | 15.6% | 16.9% | 14.5% | 1.2 | 1991 | Urban and rural | National survey of hypertension prevalence in China | 26612877 |
|  | 8125 | 16.2% | 17.4% | 15.1% | 1.2 | 1993 | Urban and rural |  |  |
|  | 8565 | 19.2% | 21.5% | 17.0% | 1.3 | 1997 | Urban and rural |  |  |
|  | 9396 | 19.5% | 21.5% | 17.5% | 1.2 | 2000 | Urban and rural |  |  |
|  | 9138 | 20.1% | 22.6% | 17.7% | 1.3 | 2004 | Urban and rural |  |  |
|  | 9163 | 18.6% | 21.1% | 16.3% | 1.3 | 2006 | Urban and rural |  |  |
|  | 9474 | 22.9% | 26.1% | 19.9% | 1.3 | 2009 | Urban and rural |  |  |
|  | 12459 | 20.9% | 23.4% | 18.7% | 1.3 | 2011 | Urban and rural |  |  |
|  | 451755 | 27.9% | 24.5% | 21.9% | 1.1 | 2012-2015 | Urban and rural | The China Hypertension Survey | 29449338 |
|  | 19477 | 27.5% | 30.8% | 24.2% | 1.3 | 2018 | Urban and rural | The China Chronic Disease and Nutrition Surveillance (CCDNS) surveys | 33271077 |
|  | 3513 | 34.10% | 40.00% | 30.70% | 1.3 | 2015-2019 | Urban and rural | NSPT cohort | / |
|  | 4092 | 19.0% | 29.0% | 9.9% | 2.9 | 2015-2021 | Urban,  high educational professional | CAS cohort | / |

This table presents the prevalence of obesity and hypertension (HTN) in adult populations across various years and regions in China. The data includes the total prevalence as well as the male and female prevalence, with the male/female ratio indicated for each dataset. The years of investigation range from 1991 to 2021, with different cohorts surveyed, including urban, rural, and high-education professional populations. Data sources include the China Health and Nutrition Survey (CHNS), the China Chronic Disease and Nutrition Surveillance (CCDNS), the National Survey of Hypertension Prevalence in China, and others. For studies not explicitly referenced, the respective cohort names (e.g., NSPT cohort, CAS cohort) are provided.

#### Table S4 Comparison of metabolic syndrome indicators between males and females in CAS and NSPT cohort

|  | **CAS cohort** | | | |  | **NSPT cohort** | | | |  | ***P* for male in CAS vs. NSPT** | ***P* for female in CAS vs. NSPT** |
| --- | --- | --- | --- | --- | --- | --- | --- | --- | --- | --- | --- | --- |
| **Metabolic syndrome indicators** | **Total** | **Male** | **Female** | ***P*** |  | **Total** | **Male** | **Female** | ***P*** |  |  |  |
| **TC, mean±sd** | 5.19±0.99 | 5.23±1.01 | 5.13±0.96 | 0.10 |  | 5.05±1.06 | 5.05±1.08 | 5.05±1.05 | 0.96 |  | 6.90E-14 | 5.73E-04 |
| **TG, mean±sd** | 1.41±1.00 | 1.67±1.15 | 1.02±0.51 | 2.15E-26 |  | 1.42±1.11 | 1.61±1.33 | 1.31±0.94 | 4.85E-12 |  | 0.04 | 5.37E-59 |
| **HDL, mean±sd** | 1.38±0.31 | 1.27±0.25 | 1.54±0.32 | 1.16E-49 |  | 1.41±0.44 | 1.32±0.40 | 1.46±0.46 | 7.52E-21 |  | 1.53E-10 | 5.50E-18 |
| **LDL, mean±sd** | 3.01±0.77 | 3.14±0.77 | 2.82±0.74 | 5.82E-12 |  | 2.89±0.93 | 2.92±0.94 | 2.87±0.93 | 0.13 |  | 3.60E-28 | 0.01 |
| **FBG, mean±sd** | 5.04±0.87 | 5.15±1.03 | 4.88±0.54 | 4.82E-07 |  | 5.43±0.95 | 5.51±0.99 | 5.38±0.93 | 2.00E-04 |  | 1.75E-53 | 5.77E-165 |

The table presents metabolic syndrome indicators, including total cholesterol (TC), triglycerides (TG), high-density lipoprotein (HDL), low-density lipoprotein (LDL), and fasting blood glucose (FBG), as mean ± standard deviation (SD) for the CAS and NSPT cohorts, stratified by gender. P-values within the CAS and NSPT cohorts reflect differences between males and females, while P-values between cohorts compare metabolic indicators for males and females in the CAS cohort versus their counterparts in the NSPT cohort.

#### Table S5 Comparison of lifestyle factors between males and females in validation set

|  | **CAS1k cohort** | | | |
| --- | --- | --- | --- | --- |
|  | **Total** | **Male** | **Female** | ***P*** |
| **lifestyle factors** |  |  |  |  |
| BMI between 18.5-23.9kg/m², n (%) | 506(47.25%) | 220(34.43%) | 286(66.20%) | 3.12E-24 |
| WC <85 cm for males and <80 cm for females, n (%) | 548(51.17%) | 227(35.52%) | 321(74.31%) | 2.84E-35 |
| No current smoking, n (%) | 947(88.42%) | 518(81.06%) | 429(99.31%) | 1.36E-19 |
| No excessive drinking, n (%) | 1019(95.14%) | 592(92.64%) | 427(98.84%) | 7.30E-06 |
| Regular physical activity, n (%) | 272(25.40%) | 179(28.01%) | 93(21.53%) | 0.02 |
| Healthy eating habits, n (%) | 162(15.13%) | 89(13.93%) | 73(16.90%) | 0.21 |

Values are presented as n (%) for lifestyle factors. P are provided for comparisons between males and females. Significant differences are indicated by P < 0.05.

#### Table S6 Description of the GWAS summary data for the PRS construction

| **Trait** | **Ancestry** | **Origin** | **Reference** | **N** | **M1** | **M2** | **M3** | **h^2^(se)** |
| --- | --- | --- | --- | --- | --- | --- | --- | --- |
| BMI | European | UK Biobank | UKBB: Pcode 21001 | 457824 | 12007571 | 11013786 | 3169089 | 0.2637 (0.0096) |
| BMI | East Asian | Biobank Japan | BBJ; Akiyama 2017 Nat Genet | 158284 | 5961600 | 5644669 | 3169089 | 0.1681 (0.0093) |
| DBP | European | UK Biobank | UKBB: Pcode 4079 | 422771 | 12008250 | 11014471 | 3169262 | 0.2044 (0.0083) |
| DBP | East Asian | Biobank Japan | BBJ; Kanai 2018 Nat Genet | 136615 | 5961600 | 5644699 | 3169262 | 0.0595 (0.0068) |
| SBP | European | UK Biobank | UKBB: Pcode 4080 | 422771 | 12008250 | 11014471 | 3169262 | 0.2058 (0.0076) |
| SBP | East Asian | Biobank Japan | BBJ; Kanai 2018 Nat Genet | 136597 | 5961600 | 5644699 | 3169262 | 0.0755 (0.0085) |

We utilized GWAS summary statistics derived from both the UK Biobank of European ancestry and BioBank Japan of East Asian ancestry for the traits of body mass index (BMI), diastolic blood pressure (DBP) and systolic blood pressure (SBP). The count of individuals in each GWAS file is indicated in the column labeled "N". Additionally, the original count of SNPs, the QC-ed count of SNPs, and the count of SNPs common to QC-ed UKBB GWAS summary, QC-ed BBJ GWAS summary, and the QC-ed CAS cohort are presented in the columns labeled "M1", “M2”, and “M3”, respectively. The SNP heritability and standard error (h^2^(se)) from each GWAS summary statistics (SNPs common to QC-ed UKBB GWAS summary, QC-ed BBJ GWAS summary, and the QC-ed CAS cohort) was estimated using LD score regression (LDSC).

#### Table S7 Fine tuning of the hyper-parameters conducted for C+T in PRS tuning set

Please refer to the MS Excel file entitled Supplementary Information.xlsx.

#### Table S8 Fine tuning of the hyper-parameters conducted for ldpred2 in PRS tuning set

Please refer to the MS Excel file entitled Supplementary Information.xlsx.

#### Table S9 Fine tuning of the hyper-parameters conducted for lassosum in PRS tuning set

Please refer to the MS Excel file entitled Supplementary Information.xlsx.

#### Table S10 Fine tuning of the hyper-parameters conducted for CT-SLEB in PRS tuning set

Please refer to the MS Excel file entitled Supplementary Information.xlsx

#### Table S1**1** Number of SNPs or CpG sites used in different PRS and MRS methods for BMI, DBP, and SBP

|  |  | **Number of SNPs in different PRS method** | | | | | | | | | | **Number of CpGs in different MRS method** | | |
| --- | --- | --- | --- | --- | --- | --- | --- | --- | --- | --- | --- | --- | --- | --- |
|  |  | **C+T** | **SCT** | **ldpred2** | **lassosum** | **PRScs** | **PRScsx** | **CT-SLEB** | **PolyPredP+** | **JointPRS** | **PROSPER** | **linear regression1** | **linear regression2** | **lasso regression** |
| BMI | UKBB | 2707939 | 2707939 | 536767 | 536767 | 574556 | 586298 | 1280441 | 3169089 | 585210 | 478775 | 1506 | 390 | 1145 |
|  | BBJ | 2707939 | 2707939 | 607362 | 607362 | 585210 |  |  |  |  |  |  |  |  |
| DBP | UKBB | 2708084 | 2708084 | 536767 | 536767 | 574555 | 586317 | 1284582 | 3146227 | 585229 | 402282 | 77 | 15 | 47 |
|  | BBJ | 2707939 | 2707939 | 607370 | 607370 | 585229 |  |  |  |  |  |  |  |  |
| SBP | UKBB | 2708084 | 2708084 | 536767 | 536767 | 574555 | 586317 | 1300934 | 3146227 | 585229 | 300211 | 107 | 18 | 97 |
|  | BBJ | 2708084 | 2708084 | 607370 | 607370 | 585229 |  |  |  |  |  |  |  |  |

The table presents the number of SNPs included in the analysis for each phenotype (BMI, DBP, and SBP) based on different ancestry GWAS datasets (UKBB or BBJ) and various PRS methods (C+T, SCT, ldpred2, lassosum, PRScs, PRScsx, CT-SLEB, PolyPred+, JointPRS, and PROSPER). Additionally, the number of CpGs included in the analysis using three MRS methods (linear regression 1, linear regression 2, and lasso regression) is also shown.

#### **Table S12 Performance of different PRS in predicting quantitative traits in tunning, testing and validation sets**

|  |  |  | **Tuning set (n = 2,030)** | | | **Testing set (n = 991)** | | | **Validation set (n = 1071)** | | |
| --- | --- | --- | --- | --- | --- | --- | --- | --- | --- | --- | --- |
| **Trait** | **Method** | **GWAS source** | **R^2^ (%)** | **95% CI Lower** | **95% CI Upper** | **R^2^ (%)** | **95% CI Lower** | **95% CI Upper** | **R^2^ (%)** | **95% CI Lower** | **95% CI Upper** |
| BMI | C+T | BBJ | 3.23 | 1.69 | 4.68 | 1.77 | 1.58 | 3.31 | 3.79 | 1.58 | 5.94 |
| BMI | C+T | UKBB | 2.83 | 1.49 | 4.11 | 4.25 | 1.53 | 6.60 | 4.36 | 1.92 | 6.52 |
| BMI | SCT | BBJ | 7.66 | 5.50 | 9.68 | 0.78 | -0.44 | 1.77 | 3.66 | 1.41 | 5.67 |
| BMI | SCT | UKBB | 10.24 | 7.78 | 12.62 | 3.38 | 0.97 | 5.52 | 1.69 | 0.11 | 3.15 |
| BMI | PRScs | BBJ | 3.29 | 1.73 | 4.72 | 3.33 | 1.00 | 5.44 | 6.36 | 3.49 | 9.07 |
| BMI | PRScs | UKBB | 2.99 | 1.50 | 4.37 | 5.56 | 2.62 | 8.17 | 6.28 | 3.50 | 8.94 |
| BMI | ldpred2 | BBJ | 3.43 | 1.88 | 4.93 | 2.46 | 0.46 | 4.34 | 4.27 | 2.05 | 6.57 |
| BMI | ldpred2 | UKBB | 1.77 | 0.56 | 2.92 | 1.75 | 0.04 | 3.22 | 0.67 | -0.34 | 1.54 |
| BMI | lassosum | BBJ | 3.10 | 1.71 | 4.45 | 2.09 | 0.23 | 3.76 | 3.55 | 1.48 | 5.60 |
| BMI | lassosum | UKBB | 1.37 | 0.19 | 2.39 | 1.75 | 0.15 | 3.19 | 0.54 | -0.47 | 1.33 |
| BMI | PRScsx | Multi-ancestry | 4.50 | 2.76 | 6.08 | 6.31 | 3.29 | 9.09 | 9.81 | 6.50 | 12.81 |
| BMI | CT-SLEB | Multi-ancestry | 5.92 | 3.90 | 7.81 | 5.52 | 2.81 | 8.26 | 6.32 | 3.55 | 8.98 |
| BMI | PolyPredP+ | Multi-ancestry | 4.42 | 2.66 | 6.04 | 5.85 | 2.91 | 8.68 | 8.85 | 5.52 | 12.05 |
| BMI | JointPRS | Multi-ancestry | 5.11 | 3.23 | 6.81 | 6.04 | 3.05 | 8.90 | 8.74 | 5.61 | 11.85 |
| BMI | PROSPER | Multi-ancestry | 5.31 | 3.28 | 7.32 | 6.17 | 3.08 | 8.98 | 7.23 | 4.20 | 9.94 |
| DBP | C+T | BBJ | 1.64 | 0.43 | 2.74 | 0.74 | -0.42 | 1.72 | 1.10 | -0.26 | 2.31 |
| DBP | C+T | UKBB | 1.95 | 0.84 | 3.02 | 2.13 | 0.28 | 3.88 | 3.17 | 0.98 | 5.18 |
| DBP | SCT | BBJ | 4.82 | 2.95 | 6.59 | 0.36 | -0.51 | 1.03 | 0.74 | -0.40 | 1.72 |
| DBP | SCT | UKBB | 6.38 | 4.34 | 8.43 | 1.13 | -0.30 | 2.31 | 2.37 | 0.51 | 4.03 |
| DBP | PRScs | BBJ | 0.94 | 0.05 | 1.77 | 1.37 | -0.14 | 2.66 | 1.60 | -0.08 | 3.07 |
| DBP | PRScs | UKBB | 2.66 | 1.18 | 4.00 | 3.16 | 0.86 | 5.34 | 3.28 | 1.02 | 5.30 |
| DBP | ldpred2 | BBJ | 1.19 | 0.18 | 2.12 | 1.53 | -0.08 | 2.98 | 2.07 | 0.33 | 3.65 |
| DBP | ldpred2 | UKBB | 1.05 | 0.06 | 1.91 | 0.81 | -0.60 | 1.98 | 0.45 | -0.46 | 1.22 |
| DBP | lassosum | BBJ | 1.35 | 0.31 | 2.34 | 1.17 | -0.28 | 2.44 | 3.08 | 0.80 | 5.14 |
| DBP | lassosum | UKBB | 1.07 | 0.14 | 1.93 | 0.61 | -0.52 | 1.56 | 0.72 | -0.47 | 1.69 |
| DBP | PRScsx | Multi-ancestry | 2.77 | 1.25 | 4.19 | 3.75 | 1.26 | 6.05 | 4.40 | 2.07 | 6.74 |
| DBP | CT-SLEB | Multi-ancestry | 10.53 | 8.08 | 12.94 | 2.71 | 0.61 | 4.58 | 4.87 | 2.34 | 7.30 |
| DBP | PolyPredP+ | Multi-ancestry | 2.74 | 1.22 | 4.17 | 3.77 | 1.52 | 6.01 | 3.60 | 1.32 | 5.84 |
| DBP | JointPRS | Multi-ancestry | 2.27 | 0.89 | 3.53 | 2.84 | 0.67 | 4.83 | 3.67 | 1.40 | 5.77 |
| DBP | PROSPER | Multi-ancestry | 3.66 | 2.01 | 5.22 | 4.03 | 1.54 | 6.34 | 5.18 | 2.54 | 7.72 |
| SBP | C+T | BBJ | 1.64 | 0.43 | 2.71 | 1.36 | -0.22 | 2.76 | 0.21 | -0.47 | 0.69 |
| SBP | C+T | UKBB | 2.03 | 0.80 | 3.21 | 1.52 | -0.20 | 3.08 | 1.29 | -0.15 | 2.52 |
| SBP | SCT | BBJ | 5.02 | 3.24 | 6.75 | 1.19 | -0.37 | 2.46 | 0.68 | -0.38 | 1.61 |
| SBP | SCT | UKBB | 7.82 | 5.63 | 9.95 | 0.74 | -0.53 | 1.76 | 1.16 | -0.22 | 2.43 |
| SBP | PRScs | BBJ | 1.19 | 0.23 | 2.03 | 1.05 | -0.48 | 2.31 | 0.47 | -0.46 | 1.21 |
| SBP | PRScs | UKBB | 2.24 | 0.92 | 3.37 | 3.85 | 1.35 | 6.28 | 2.23 | 0.24 | 4.01 |
| SBP | ldpred2 | BBJ | 1.58 | 0.48 | 2.52 | 1.13 | -0.39 | 2.37 | 0.51 | -0.40 | 1.28 |
| SBP | ldpred2 | UKBB | 1.15 | 0.09 | 2.07 | 0.91 | -0.45 | 2.11 | 0.14 | -0.48 | 0.55 |
| SBP | lassosum | BBJ | 1.61 | 0.43 | 2.71 | 1.37 | -0.32 | 2.81 | 0.91 | -0.33 | 1.94 |
| SBP | lassosum | UKBB | 1.03 | 0.09 | 1.84 | 0.99 | -0.33 | 2.19 | 0.06 | -0.43 | 0.36 |
| SBP | PRScsx | Multi-ancestry | 2.69 | 1.32 | 4.01 | 4.23 | 1.40 | 6.75 | 2.40 | 0.55 | 4.10 |
| SBP | CT-SLEB | Multi-ancestry | 5.40 | 3.53 | 7.18 | 3.21 | 0.77 | 5.44 | 2.37 | 0.60 | 3.98 |
| SBP | PolyPredP+ | Multi-ancestry | 2.59 | 1.24 | 3.83 | 3.47 | 1.01 | 5.80 | 1.78 | 0.23 | 3.25 |
| SBP | JointPRS | Multi-ancestry | 2.56 | 1.30 | 3.80 | 3.40 | 0.84 | 5.79 | 1.62 | 0.16 | 2.99 |
| SBP | PROSPER | Multi-ancestry | 3.22 | 1.70 | 4.68 | 4.41 | 1.50 | 7.12 | 2.52 | 0.52 | 4.24 |

The table summarizes the R2 values and their 95% confidence intervals (CI) for BMI, DBP and SBP across three datasets: tuning set (n=2,030), testing set (n=991), and validation set (n=1,071). Results are shown for various PRS methods, including C+T, SCT, PRScs, ldpred2, lassosum, PRScsx, CT-SLEB, PolyPred+, JointPRS, and PROSPER. The GWAS used include UKBB, BBJ, and multi-ancestry datasets (both of all). The R2 values represent the proportion of variance explained by the PRS for residuals from the regression of each trait on age, sex, and six genomic principal components. Confidence intervals were estimated using a bootstrap resampling approach.

#### **Table S13 The performance of PRSs in PGS Catalog**

|  |  | **Testing set (*n* = 991)** | | | **Validation set (*n* = 1071)** | | |
| --- | --- | --- | --- | --- | --- | --- | --- |
| **Method** | **trait** | **R^2^ (%)** | **95% CI Lower** | **95% CI Upper** | **R^2^ (%)** | **95% CI Lower** | **95% CI Upper** |
| **PGS000298** | **BMI** | 1.87% | -0.07% | 3.61% | 3.73% | 1.19% | 6.00% |
| **PGS000770** | **BMI** | 0.57% | -0.46% | 1.38% | 1.21% | -0.24% | 2.50% |
| **PGS000910** | **BMI** | 3.62% | 1.21% | 5.83% | 2.75% | 0.87% | 4.44% |
| **PGS000921** | **BMI** | 3.04% | 0.59% | 5.18% | 3.34% | 1.27% | 5.32% |
| **PGS002251** | **BMI** | 0.34% | -0.61% | 1.09% | 1.46% | -0.03% | 2.80% |
| **PGS002751** | **BMI** | 0.04% | -0.38% | 0.28% | 0.00% | -0.34% | 0.17% |
| **PGS003842** | **BMI** | 0.83% | -0.50% | 1.91% | 0.31% | -0.55% | 0.97% |
| **PGS003843** | **BMI** | 1.27% | -0.30% | 2.65% | 2.47% | 0.50% | 4.33% |
| **PGS003844** | **BMI** | 0.32% | -0.41% | 0.90% | 1.05% | -0.39% | 2.28% |
| **PGS003845** | **BMI** | 0.13% | -0.47% | 0.56% | 0.43% | -0.57% | 1.22% |
| **PGS003846** | **BMI** | 0.04% | -0.40% | 0.29% | 0.01% | -0.38% | 0.21% |
| **PGS003847** | **BMI** | 0.76% | -0.33% | 1.75% | 2.13% | 0.26% | 3.79% |
| **PGS003848** | **BMI** | 1.54% | -0.06% | 2.90% | 2.46% | 0.40% | 4.34% |
| **PGS003884** | **BMI** | 2.66% | 0.62% | 4.50% | 3.42% | 1.30% | 5.41% |
| **PGS003885** | **BMI** | 0.49% | -0.47% | 1.27% | 1.44% | 0.11% | 2.63% |
| **PGS003886** | **BMI** | 4.31% | 1.75% | 6.83% | 7.50% | 4.49% | 10.22% |
| **PGS003887** | **BMI** | 4.43% | 1.83% | 6.90% | 6.38% | 3.77% | 9.00% |
| **PGS003980** | **BMI** | 1.69% | 0.05% | 3.19% | 2.92% | 0.90% | 4.79% |
| **PGS003996** | **BMI** | 2.85% | 0.63% | 4.81% | 3.25% | 1.18% | 5.21% |
| **PGS004012** | **BMI** | 2.61% | 0.41% | 4.52% | 2.43% | 0.54% | 4.17% |
| **PGS004022** | **BMI** | 1.44% | -0.16% | 2.84% | 2.74% | 0.78% | 4.58% |
| **PGS004037** | **BMI** | 2.11% | 0.09% | 3.84% | 2.28% | 0.40% | 3.97% |
| **PGS004050** | **BMI** | 1.21% | -0.30% | 2.50% | 2.42% | 0.65% | 4.10% |
| **PGS004066** | **BMI** | 3.07% | 0.75% | 5.26% | 3.63% | 1.38% | 5.66% |
| **PGS004080** | **BMI** | 2.60% | 0.57% | 4.43% | 3.91% | 1.50% | 6.10% |
| **PGS004096** | **BMI** | 2.46% | 0.43% | 4.33% | 2.33% | 0.42% | 4.02% |
| **PGS004104** | **BMI** | 0.57% | -0.46% | 1.40% | 1.11% | -0.33% | 2.31% |
| **PGS004120** | **BMI** | 0.74% | -0.41% | 1.69% | 0.63% | -0.44% | 1.48% |
| **PGS004134** | **BMI** | 2.00% | 0.09% | 3.72% | 2.87% | 0.76% | 4.86% |
| **PGS004150** | **BMI** | 3.09% | 0.87% | 5.10% | 3.69% | 1.47% | 5.71% |
| **PGS004319** | **BMI** | 0.19% | -0.44% | 0.67% | 0.42% | -0.52% | 1.15% |
| **PGS004609** | **BMI** | 2.16% | 0.23% | 3.90% | 2.18% | 0.33% | 3.87% |
| **PGS004610** | **BMI** | 1.57% | 0.05% | 2.99% | 1.38% | -0.17% | 2.74% |
| **PGS004864** | **BMI** | 0.03% | -0.36% | 0.24% | 0.22% | -0.46% | 0.72% |
| **PGS004902** | **BMI** | 4.74% | 2.03% | 7.39% | 5.67% | 2.88% | 8.35% |
| **PGS004982** | **BMI** | 0.80% | -0.24% | 1.74% | 1.90% | 0.02% | 3.53% |
| **PGS004983** | **BMI** | 0.16% | -0.47% | 0.62% | 0.89% | -0.35% | 1.96% |
| **PGS004984** | **BMI** | 1.41% | -0.26% | 2.89% | 2.41% | 0.49% | 4.28% |
| **PGS004985** | **BMI** | 4.75% | 2.00% | 7.17% | 5.34% | 2.35% | 8.13% |
| **PGS004986** | **BMI** | 4.39% | 1.72% | 6.86% | 5.79% | 2.78% | 8.55% |
| **PGS004987** | **BMI** | 3.65% | 1.25% | 5.76% | 4.84% | 2.31% | 7.25% |
| **PGS004988** | **BMI** | 0.78% | -0.32% | 1.73% | 1.34% | -0.22% | 2.69% |
| **PGS004989** | **BMI** | 3.99% | 1.52% | 6.33% | 4.29% | 1.89% | 6.69% |
| **PGS004990** | **BMI** | 1.51% | -0.21% | 3.05% | 1.64% | 0.00% | 3.05% |
| **PGS004991** | **BMI** | 3.26% | 0.82% | 5.46% | 4.20% | 1.63% | 6.46% |
| **PGS004992** | **BMI** | 4.75% | 1.99% | 7.28% | 5.99% | 3.14% | 8.71% |
| **PGS004993** | **BMI** | 3.75% | 1.43% | 5.97% | 5.89% | 3.01% | 8.57% |
| **PGS004994** | **BMI** | 5.60% | 2.84% | 8.19% | 5.32% | 2.62% | 7.83% |
| **PGS000302** | **DBP** | 1.97% | 0.18% | 3.59% | 1.55% | 0.08% | 2.94% |
| **PGS002258** | **DBP** | 1.40% | -0.14% | 2.74% | 1.57% | 0.02% | 2.97% |
| **PGS003964** | **DBP** | 4.90% | 2.17% | 7.41% | 2.62% | 0.86% | 4.30% |
| **PGS004232** | **DBP** | 1.03% | -0.34% | 2.18% | 0.70% | -0.41% | 1.65% |
| **PGS000301** | **SBP** | 2.41% | 0.46% | 4.22% | 1.56% | -0.01% | 2.93% |
| **PGS002257** | **SBP** | 2.14% | 0.37% | 3.81% | 1.62% | 0.01% | 2.99% |
| **PGS002275** | **SBP** | 1.51% | -0.12% | 2.94% | 0.90% | -0.37% | 1.93% |
| **PGS003588** | **SBP** | 1.37% | -0.18% | 2.69% | 2.24% | 0.48% | 3.87% |
| **PGS003882** | **SBP** | 3.51% | 1.01% | 5.76% | 2.64% | 0.75% | 4.48% |
| **PGS003968** | **SBP** | 4.37% | 1.72% | 6.92% | 2.19% | 0.41% | 3.86% |
| **PGS004231** | **SBP** | 1.82% | 0.03% | 3.43% | 1.04% | -0.29% | 2.17% |
| **PGS005008** | **SBP** | 0.52% | -0.47% | 1.32% | 0.90% | -0.34% | 1.93% |
| **PGS005009** | **SBP** | 0.32% | -0.53% | 0.99% | 0.75% | -0.44% | 1.72% |
| **PGS005010** | **SBP** | 1.61% | -0.16% | 3.21% | 0.35% | -0.44% | 0.97% |
| **PGS005011** | **SBP** | 1.98% | 0.12% | 3.63% | 1.88% | 0.24% | 3.37% |
| **PGS005012** | **SBP** | 2.07% | 0.07% | 3.89% | 1.97% | 0.22% | 3.51% |
| **PGS005013** | **SBP** | 1.79% | 0.08% | 3.39% | 1.98% | 0.18% | 3.61% |
| **PGS005014** | **SBP** | 0.30% | -0.48% | 0.90% | 0.54% | -0.46% | 1.40% |
| **PGS005015** | **SBP** | 2.09% | 0.10% | 3.94% | 2.50% | 0.50% | 4.24% |
| **PGS005016** | **SBP** | 1.26% | -0.24% | 2.52% | 0.56% | -0.45% | 1.43% |
| **PGS005017** | **SBP** | 1.59% | -0.03% | 3.06% | 2.14% | 0.27% | 3.77% |
| **PGS005018** | **SBP** | 1.75% | -0.11% | 3.35% | 2.17% | 0.32% | 3.78% |
| **PGS005019** | **SBP** | 2.19% | 0.12% | 4.03% | 1.78% | 0.14% | 3.29% |
| **PGS005020** | **SBP** | 2.46% | 0.44% | 4.37% | 2.25% | 0.42% | 3.93% |

All PRSs for BMI, DBP, and SBP selected from the PGS Catalog were analyzed in the PRS testing set (n=991) and the validation set (n=1,071). Each trait was regressed on age, sex, and six genomic principal components, with the residuals from this regression used as the dependent variable. R-squared (R2) and 95% confidence intervals were estimated using a 10,000 bootstrap resampling approach.

#### **Table S14 Peformance of different MRS in tunning, testing and validaltion sets**

|  |  | **Tuning set (n=2030)** | | | **Testing set (n=991)** | | | **Validation set (n=1071)** | | |
| --- | --- | --- | --- | --- | --- | --- | --- | --- | --- | --- |
| **Trait** | **MRS method** | **R**^2^ **(%)** | **95% CI Lower** | **95% CI Upper** | **R**^2^ **(%)** | **95% CI Lower** | **95% CI Upper** | **R**^2^ **(%)** | **95% CI Lower** | **95% CI Upper** |
| BMI | lasso | 72.10% | 70.02% | 74.28% | 8.48% | 6.20% | 11.53% | 10.03% | 6.68% | 12.78% |
|  | linear 1 | 13.50% | 10.90% | 16.21% | 5.98% | 3.05% | 8.46% | 5.75% | 3.09% | 8.39% |
|  | linear 2 | 13.40% | 10.77% | 16.13% | 6.64% | 3.98% | 8.87% | 5.13% | 2.72% | 7.44% |
| DBP | lasso | 4.07% | 2.32% | 5.57% | 1.61% | 0.34% | 2.94% | 4.68% | 2.22% | 7.18% |
|  | linear 1 | 0.88% | -0.10% | 1.76% | 0.64% | -0.17% | 1.22% | 2.66% | 0.40% | 4.60% |
|  | linear 2 | 1.10% | 0.36% | 1.95% | 0.70% | -0.46% | 1.57% | 2.32% | 0.35% | 4.29% |
| SBP | lasso | 9.11% | 6.54% | 11.67% | 3.02% | 1.43% | 4.92% | 3.70% | 1.30% | 5.99% |
|  | linear 1 | 2.81% | 1.37% | 4.29% | 1.64% | 0.48% | 2.78% | 2.10% | 0.51% | 3.65% |
|  | linear 2 | 3.02% | 1.68% | 4.21% | 2.13% | 0.76% | 3.59% | 3.40% | 1.07% | 5.70% |

For each trait (BMI, DBP, and SBP), PRS were constructed using different methods: lasso and two linear modeling approaches. Linear 1 refers to linear modeling using all pre-selected CpGs, while linear 2 refers to linear modeling considering only those CpGs with an association p-value < 0.05 in the data. Lasso regression considered CpGs remained in the model. The performance of each method was assessed in three sets: the MRS tuning set (n = 2,030), MRS testing set (n = 991), and validation set (n = 1,071). R-squared (R²) values and 95% confidence intervals (CI) are reported through 10,000 bootstrap resampling for each set.

#### Table S1**5** **ORs for Obesity, HTN, and OrH across PRS and MRS in the validation set**

| **Trait** | **Risk Score** | **OR (Per SD)** | ***P*** | **Q2 vs. Q1** | **Q3 vs. Q1** | **Q4 vs. Q1** | **Q5 vs. Q1** |
| --- | --- | --- | --- | --- | --- | --- | --- |
| Obesity |  |  |  |  |  |  |  |
| OR (95% CI) | PRS_BMI | 2.13[1.77-2.57] | 1.70E-11 | 2.72[1.11-6.66] | 7.58[3.35-17.14] | 5.59[2.44-12.78] | 12.63[5.67-28.13] |
| OR (95% CI) | MRS_BMI | 1.88[1.57-2.24] | 4.94E-09 | 1.34[0.64-2.81] | 2.36[1.20-4.65] | 2.67[1.37-5.19] | 5.77[3.07-10.86] |
| HTN |  |  |  |  |  |  |  |
| OR (95% CI) | PRS_HTN | 1.68[1.47-1.91] | 1.04E-10 | 0.80[0.51-1.24] | 1.82[1.22-2.70] | 2.53[1.71-3.76] | 2.93[1.98-4.35] |
| OR (95% CI) | MRS_HTN | 1.65[1.43-1.89] | 3.85E-09 | 1.86[1.15-3.00] | 2.18[1.36-3.48] | 2.71[1.70-4.32] | 4.02[2.53-6.38] |
| OrH |  |  |  |  |  |  |  |
| OR (95% CI) | PRS_BMI | 1.42[1.23-1.63] | 3.88E-05 | 1.08[0.68-1.72] | 1.28[0.82-2.00] | 1.67[1.09-2.58] | 2.36[1.54-3.61] |
| OR (95% CI) | PRS_HTN | 1.45[1.26-1.66] | 1.63E-05 | 0.95[0.59-1.54] | 1.98[1.28-3.05] | 2.50[1.63-3.84] | 2.02[1.30-3.15] |
| OR (95% CI) | MRS_BMI | 1.52[1.31-1.77] | 4.42E-06 | 2.18[1.25-3.81] | 3.19[1.86-5.46] | 2.62[1.53-4.48] | 6.03[3.59-10.11] |
| OR (95% CI) | MRS_HTN | 1.72[1.46-2.02] | 4.99E-08 | 1.87[0.99-3.51] | 3.04[1.67-5.55] | 3.40[1.88-6.17] | 6.52[3.64-11.66] |

The odds ratio (OR) and 95% confidence interval (CI) for obesity, HTN, and OrH associated with each standard deviation increment in the selected risk scores (PRS and MRS) are presented. Additionally, the OR (compared to the first quantile, Q1) and 95% CI are reported across quintiles (Q2–Q5) for each disease. All analyses were adjusted for age and sex in the validation set (n = 1,071).

#### **Table S16 Comparison of metabolic and lifestyle factors among participants stratified by prediction accuracy of MRS for BMI, DBP, and SBP**

| **Trait** | **Trait** | **Group 1: MRS predictions underestimated the observed values** | **Group 2: MRS predictions closely matched the observed values** | **Group 3: MRS predictions overestimated the observed values** | **P(Group 1 vs. Group 2)** | **P(Group 3 vs. Group 2)** |
| --- | --- | --- | --- | --- | --- | --- |
| BMI | TC, mmol/L | 5.13±0.99 | 5.20±1.00 | 5.21±0.96 | 0.29 | 0.94 |
|  | TG, mmol/L | 1.26±0.76 | 1.39±1.03 | 1.61±1.08 | 0.04 | 0.01 |
|  | HDL, mmol/L | 1.47±0.36 | 1.39±0.30 | 1.26±0.24 | 3.94E-03 | 5.69E-10 |
|  | LDL, mmol/L | 2.88±0.77 | 3.01±0.77 | 3.13±0.76 | 0.03 | 0.05 |
|  | FBG, mmol/L | 5.01±0.98 | 5.02±0.83 | 5.15±0.88 | 0.86 | 0.06 |
|  | Healthy life style | 3.90±0.93 | 3.29±1.27 | 2.41±0.82 | 3.67E-13 | 1.81E-28 |
| DBP | TC, mmol/L | 5.13±0.96 | 5.16±0.98 | 5.35±1.04 | 0.62 | 0.02 |
|  | TG, mmol/L | 1.35±0.78 | 1.40±1.11 | 1.49±0.81 | 0.50 | 0.19 |
|  | HDL, mmol/L | 1.36±0.30 | 1.39±0.31 | 1.35±0.32 | 0.14 | 0.06 |
|  | LDL, mmol/L | 2.97±0.74 | 2.97±0.76 | 3.18±0.83 | 0.91 | 1.10E-03 |
|  | FBG, mmol/L | 5.04±1.19 | 5.01±0.78 | 5.16±0.75 | 0.73 | 0.01 |
|  | Healthy life style | 3.49±1.17 | 3.24±1.24 | 2.96±1.17 | 0.01 | 2.76E-03 |
| SBP | TC, mmol/L | 5.13±0.95 | 5.16±0.95 | 5.36±1.12 | 0.68 | 0.02 |
|  | TG, mmol/L | 1.34±0.75 | 1.40±0.95 | 1.50±1.30 | 0.33 | 0.30 |
|  | HDL, mmol/L | 1.37±0.30 | 1.38±0.31 | 1.39±0.32 | 0.80 | 0.68 |
|  | LDL, mmol/L | 2.96±0.75 | 2.99±0.75 | 3.14±0.85 | 0.66 | 0.02 |
|  | FBG, mmol/L | 4.92±0.69 | 5.04±0.92 | 5.16±0.90 | 0.03 | 0.10 |
|  | Healthy life style | 3.36±1.24 | 3.23±1.25 | 3.12±1.11 | 0.18 | 0.20 |

Data are presented as mean±standard deviation. P-values indicate the statistical significance of differences between Group 1 vs. Group 2 and Group 3 vs. Group 2.

Healthy lifestyle: Composite lifestyle score ranging from 0 to 6, with higher scores indicating healthier lifestyle behaviors. Group 1:MRS predictions underestimated the observed values: Participants whose MRS significantly underestimated the trait (lowest 10% quantile of the difference between predicted and observed values). Group 2:MRS predictions closely matched the observed values: Participants whose MRS predictions closely matched their observed trait values (middle 80% quantiles). Group 3:MRS predictions overestimated the observed values: Participants whose MRS significantly overestimated the trait (highest 10% quantile of the difference between predicted and observed values).

### Supplemental Figures

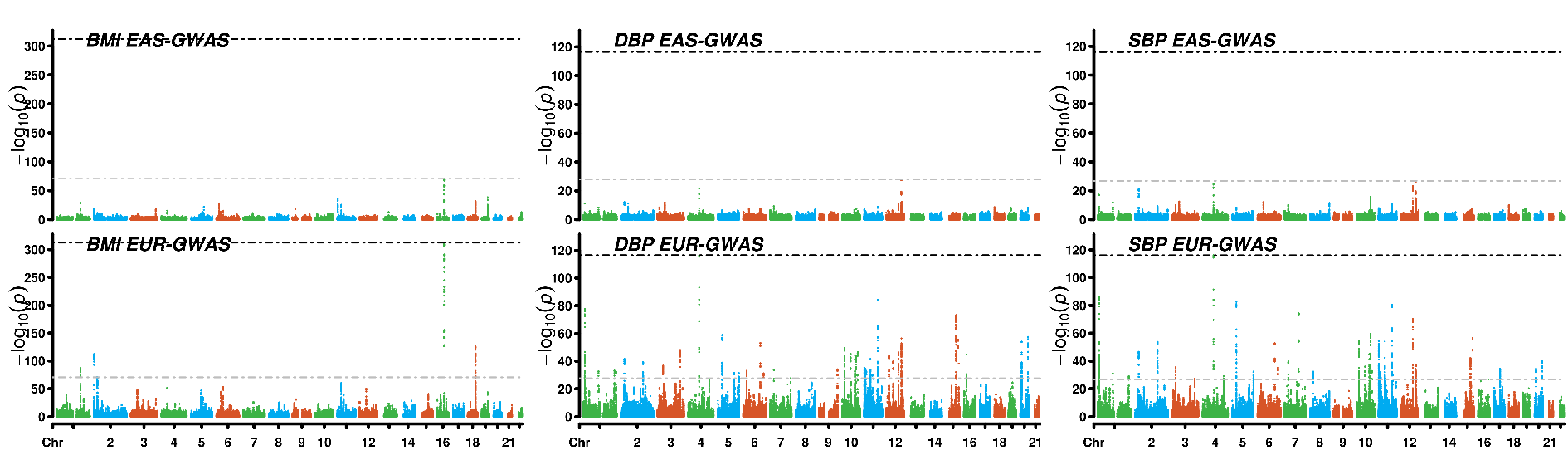

#### Figure S1 Manhattan plot of GWASs on BMI, DBP and SBP from UK Biobank and Biobank Japan

The *P* for each variant in the EAS (Biobank Japan) and EUR (UK Biobank) GWASs are depicted for body mass index (BMI), diastolic blood pressure (DBP) and systolic blood pressure (SBP). Chromosomal location is represented on the horizontal axis, and the negative log of the *P* is represented on the vertical axis. For each trait, the grey dashed lines indicate the most significant *P* in EAS GWAS and black dashed lines indicates the most significant *P* in EUR GWAS.

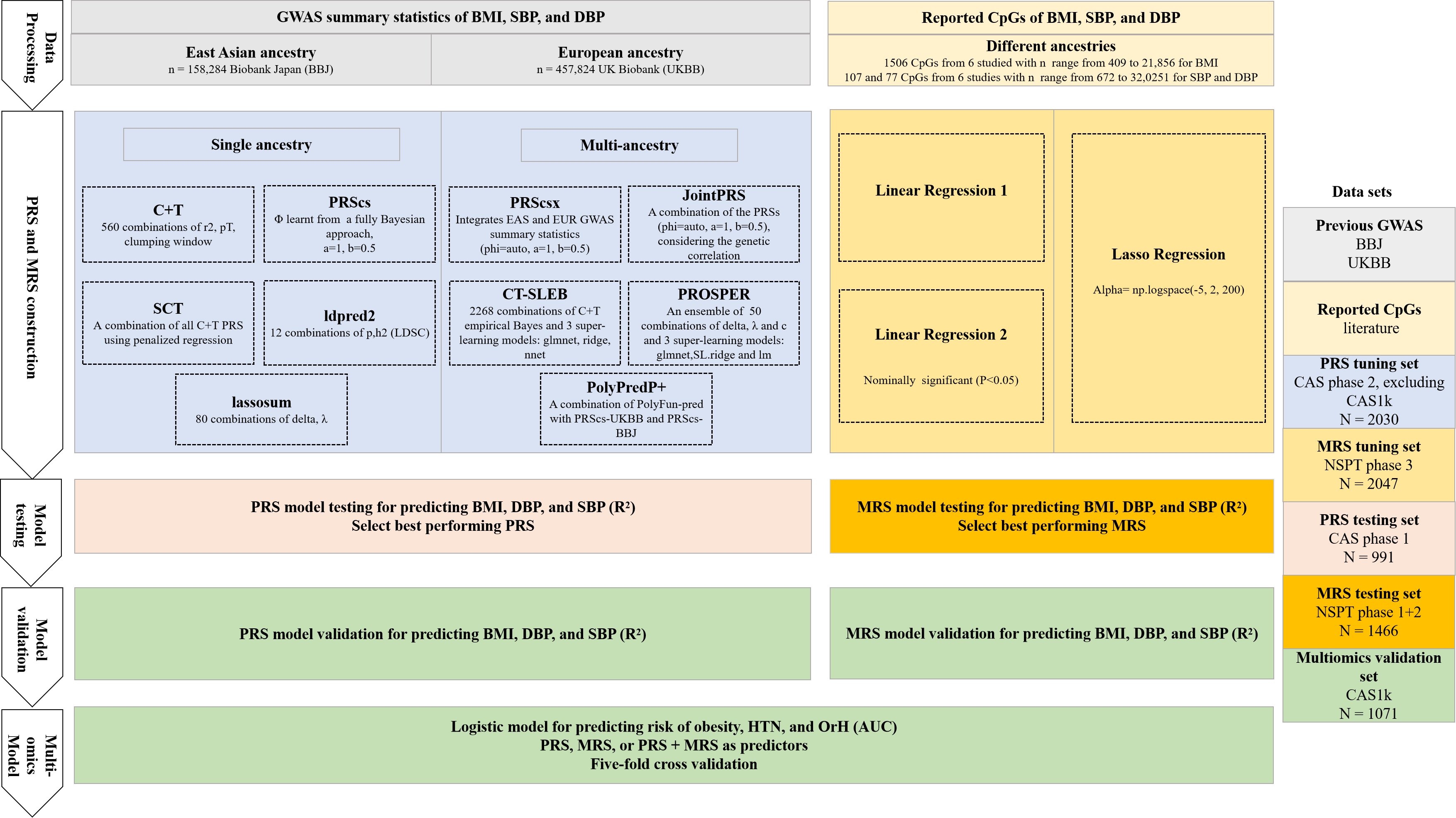

#### Figure S2 Workflow of an integrative multi-omics model combining PRS and MRS for OrH risk prediction

This study presents an integrative multi-omics model that combines PRS and MRS to enhance the profiling of OrH risk. Utilizing GWAS summary statistics from Biobank Japan (BBJ) and the UK Biobank (UKBB), we systematically evaluated various PRS methodologies, including C+T, SCT, PRScs, ldpred2, lassosum, PRScsx, CT-SLEB, PolyPred+, JointPRS, and PROSPER. A Chinese cohort of 4,092 individuals from the Chinese Academy of Sciences (CAS) was divided into three datasets: Phase 2 (excluding CAS1k) for hyperparameter tuning (n = 2,030), Phase 1 for model testing (n = 991), and CAS1k for final validation (n = 1,071). Additionally, we developed multiple MRS models (Linear Regression 1, Linear Regression 2, and Lasso Regression) based on previous EWAS findings, using data from 3,513 Chinese participants in the National Survey of Physical Traits (NSPT) cohort. NSPT Phase 3 was employed for MRS parameter tuning (n = 2,047), while NSPT Phases 1 and 2 served as testing sets (n = 1,466) to evaluate MRS performance. The predictive efficacy of the MRS for OrH risk was further validated in the CAS1k validation set (n = 1,071). After deriving the optimal parameters for linear models of PRS and MRS for BMI, SBP, and DBP, these scores were integrated into a multi-omics score. The performance of the integrated model in predicting binary outcomes—obesity, HTN, and OrH—was assessed in the CAS1k validation set (n = 1,071) using a five-fold cross-validation approach.

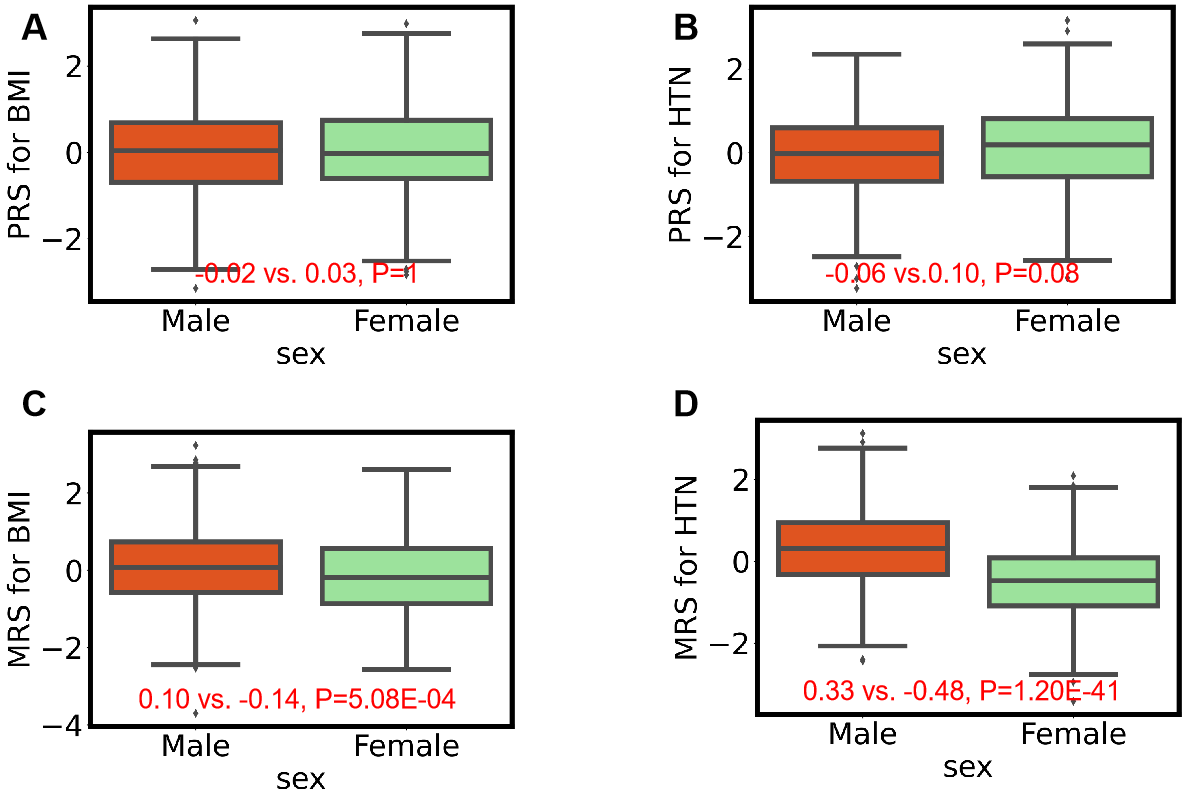

#### Figure S3 Sex-specific distributions of PRS and MRS in the validation set

Panels (A) and (B) show the distributions of PRS_BMI and PRS_HTN (the average of PRS_DBP and PRS_SBP), respectively, with no significant difference in PRS_BMI between males and females (P = 1) and a marginally non-significant difference in PRS_HTN (P = 0.08). Panels (C) and (D) present the distributions of MRS_BMI and MRS_HTN (the average of MRS_DBP and MRS_SBP), respectively, highlighting statistically significant differences between sexes for both MRS_BMI (P = 5.08E-04) and MRS_HTN (P = 1.20E-41). The analysis was conducted in the validation set (n = 1,071), and the boxplots represent the median, interquartile range, and overall data distribution for each group.
